# Interrupting the intergenerational cycle of violence: protocol for a three-generational longitudinal mixed-methods study in South Africa (INTERRUPT_VIOLENCE)

**DOI:** 10.1101/2022.11.29.22282893

**Authors:** Franziska Meinck, Nataly Woollett, Hannabeth Franchino-Olsen, Mpho Silima, Christina Thurston, Ansie Fouché, Kopano Monaisa, Nicola Christofides

## Abstract

**Introduction:** Violence is a global social and human rights issue with serious public health implications across the life-course. Interpersonal violence is transmitted across generations and there is an urgent need to understand the mechanisms of this transmission to identify and inform interventions and policies for prevention and response. We lack an evidence-base for understanding the underlying mechanisms of the intra- and intergenerational transmission of violence as well as potential for intervention, particularly in regions with high rates of interpersonal violence such as sub-Saharan Africa. The study has three aims: 1) to identify mechanisms of violence transmission across generations and by gender through quantitative and qualitative methods; 2) to examine the effect of multiple violence experience on health outcomes, victimisation and perpetration; 3) to investigate the effect of structural risk factors on violence transmission; and 4) to examine protective interventions and policies.

**Methods and analysis:** INTERRUPT_VIOLENCE is a mixed-methods three-generational longitudinal study. It builds on a two-wave existing cohort study of 1665 adolescents in South Africa interviewed in 2010/11 and 2011/12. For wave three and possible future waves, the original participants (now young adults), their oldest child (aged 6+), and their former primary caregiver will be recruited. Quantitative surveys will be carried out followed by qualitative in-depth interviews with a subset of 30 survey families. Adults will provide informed consent, while children will be invited to assent following adult consent for child participation. Stringent distress and referral protocols will be in place for the study. Triangulation will be used to deepen interpretation of findings. Qualitative data will be analysed thematically, quantitative data using advanced longitudinal modelling.

**Ethics and dissemination:** Ethical approval was granted by the University of Edinburgh, University of the Witwatersrand, North-West University, and the Provincial Department of Health Mpumalanga. Results will be published in peer-reviewed journals, at scientific meetings, and policy briefs.

**Article summary:** *Strengths and limitations of this study:*

- This is the first study on intergenerational violence transmission in a three-generational longitudinal sample in Southern Africa
- It measures not only violence against children (physical abuse, emotional abuse, sexual abuse, neglect and exposure to domestic violence), but also intimate partner violence experience and perpetration, non-partner sexual violence experience and perpetration, bullying experience and perpetration, community violence experience, witnessing and perpetration, and structural violence experience in addition to risk and protective factors on all levels of the socio-ecological model
- The study will employ a mixed-methods approach to develop the first empirically generated theoretical framework to transform our understanding of causes, effects, and prevention of violence transmission in Southern Africa
- The proposed study represents a major scientific advance in understanding the transmission and prevention of violence and will impact a critically important societal and public health challenge of our time
- While the study involves a large eligible sample, challenges with tracing and attrition 10 years after the original data collection may reduce sample size substantially

## Introduction

Violence experience and perpetration are serious social and public health issues. Eliminating all forms of violence is recognised as an important target for development within the United Nations Sustainable Development Goals (Target 5.2 and 16.2)^1^ and is a policy priority for many international non-governmental organisations such as the World Health Organization (WHO) and UNICEF [1,2]. Violence experience across the lifespan is associated with long-term poor physical, mental and reproductive health outcomes [3–6], reduced academic performance, poor social and cognitive functioning, and changes in brain development [7–9]. It is also linked with a number of high-risk behaviours such as smoking, alcohol and drug use, and sexual risk, which in turn increase risk for cancer and other non-communicable diseases and sexually transmitted infections [10–13].

The global agenda focuses on the prevention of violence against women and children. It is estimated that 1 billion children experience violence annually [14], while at least one in three women has experienced physical and/or sexual intimate partner violence (IPV) in her lifetime, with exposures starting in adolescence [15]. The majority of those experiencing violence do not experience just one type of violence at any given time but are subjected to multiple types of violence which co-occur and increase risks for poor outcomes [16].

A multitude of studies have found associations between violence experience in childhood and violence experience and perpetration in late adolescence and adulthood [17]. Violence experience is defined as first-hand experience of violence or being a victim of violence. Violence perpetration is defined as the act of carrying out a violent act against another. The two concepts are closely linked; victims can be perpetrators and perpetrators can be victims at the same time. Most of the literature focuses on male perpetration and female victimisation [18] due to broader gender and power dynamics in relationships, hegemonic masculinities [19] and large bodies of evidence demonstrating more severe perpetration of violence by men against women than the other way around [20]. However, both men and women can be victims and perpetrators, though differences in mechanisms for both genders remain understudied. Violence and adversity in childhood are considered the main risk factors for violence experience and perpetration in adulthood [21,22]. A recent systematic review has established that families in which a parent experienced childhood maltreatment are at much higher risk of maltreating their own children compared to those without maltreatment histories [23]. Similarly, children who are exposed to IPV between their parents or experience peer violence or child maltreatment are more likely to experience or perpetrate IPV during adulthood [18,24,25]. This so-called ‘intergenerational transmission of violence’ (see Figure 1) is much discussed, but there are substantial limitations in the validity and applicability globally because much of the evidence is based on studies from high-income countries using official child protection records or cross-sectional data with adults retrospectively reporting their own child maltreatment experience [17,22].

**Figure 1:**
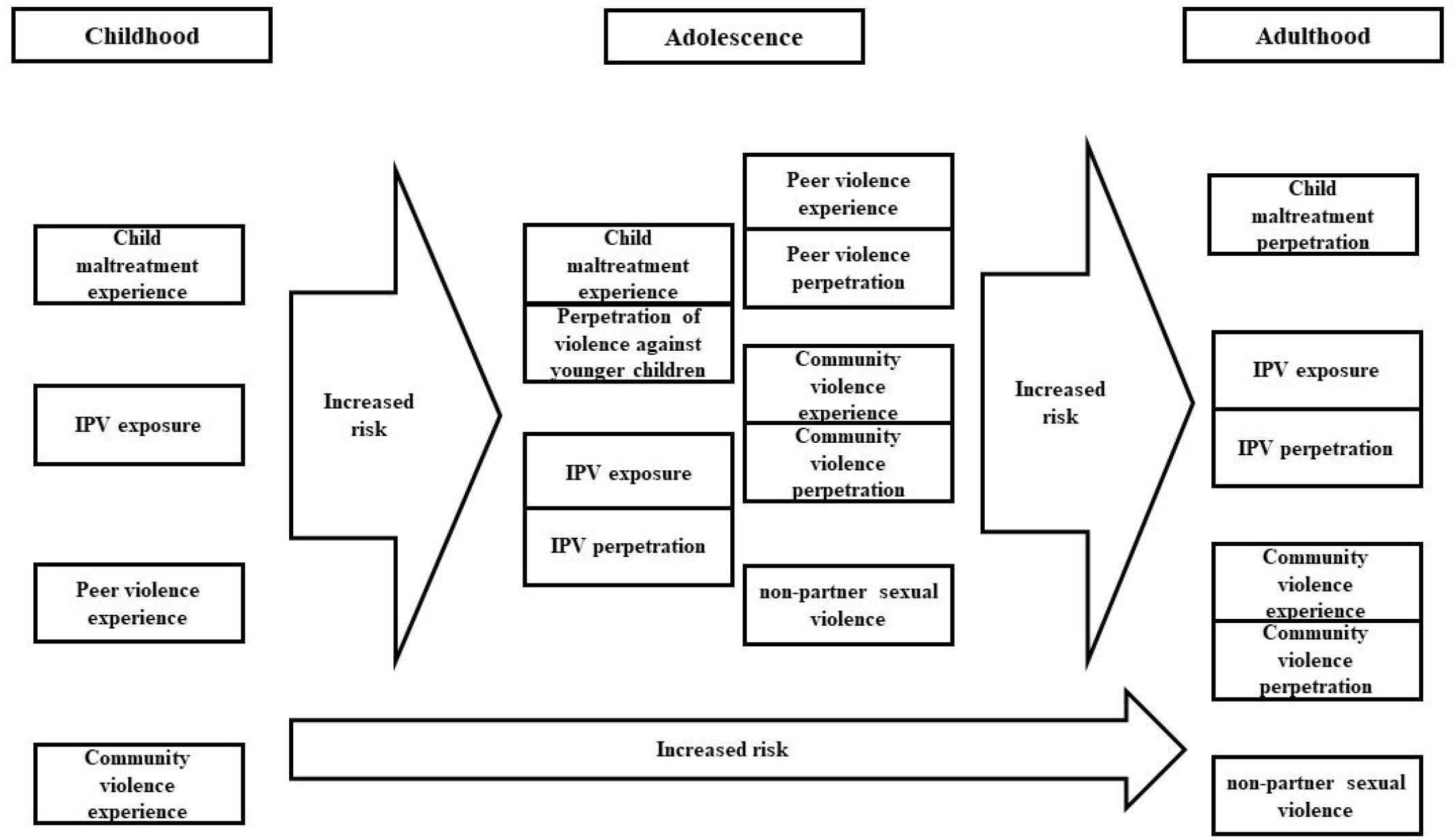
Conceptual model on the intergenerational transmission of violence independent of context.

To date, little research using longitudinal data has been conducted on the intergenerational transmission of violence, particularly in low- and middle-income countries (LMIC). Of those studies that have been conducted, most focus on one generation of respondents [26,27] or two at the most [28]. Exemplary multi-generational cohort studies in Malawi and South Africa exist, but none thus far investigate intergenerational transmission of violence [29,30]. Little is therefore known about risk pathways from childhood violence experience to adult violence experience, perpetration and parenting in contexts where multiple types of violence are prevalent. What is known is that while risk for violence perpetration is higher among adults who experienced violence in childhood, only approximately 1 in 5 maltreated children go on to perpetrate violence [22]. These findings suggest that there must be protective mechanisms for perpetration relevant to prevention which to date have not been studied in most of the global population, especially in Southern Africa. To prevent violence across the life-course, it is essential to generate a rigorous longitudinal evidence-base and to develop a theoretical framework to allow better understanding of, and protection against, violence. This will help build a foundation for future research towards designing effective policies and programmes for intervention and prevention.

### Theoretical framework for the intergenerational transmission of violence

Theoretical explanations on the driving factors of violence and the intergenerational transmission of various types of violence have been studied in multiple contexts and through different lenses. While the overarching idea of intergenerational transmission in the field of violence is well studied for intimate partner violence (IPV) and trauma, much less is known about it in terms of childhood exposures to multiple diverse violence types and victimisation and perpetration later in life. Risk factors associated and assumed to be causally affecting violence victimisation and perpetration are frequently considered in an ecological framework [31,32]. This framework posits that the interplay of risk and protective factors at individual, relationship, community, and societal levels determines one’s risk for violence experience and perpetration across the lifespan in the context of changes in the person and their environment over time [33]. At the individual level, these factors may be age, sex, education, early violence experience, disability, and poor health. At the relationship level, these factors may include exposure to violent parental conflict, parental absence, low socio-economic status, or poor parenting practices. At the community level, factors include concentrated poverty, high crime levels, epidemics which disrupt community life, deficient family caring arrangements that lead to community break down, inadequate victim care services, and weak institutional policies. At the societal level, factors include gender inequalities, poverty, legal and cultural norms that support violence, and weak safety nets [34]. Violence transmission is more likely when risk factors outweigh protective factors [32]. Research on violence victimisation and perpetration over the past decades has mostly focused on factors at the individual and relationship levels, with significantly fewer studies focusing on community and societal factors or on the ways to mitigate adverse social factors. Thus, there is an urgent need for a theoretical framework that captures the complex mechanisms that underpin the transmission of violence over time in families and communities, and examines whether factors such as poverty, poor service delivery, and ongoing epidemics (e.g. HIV) or effective social protection provision amplify or mitigate violence risk.

### Aim

The overarching aim of the proposed study is to address these significant research gaps and transform our understanding of violence transmission by conducting the first and largest longitudinal study on the intergenerational transmission of violence among young people of reproductive age in Southern Africa and to develop an empirically generated theoretical framework for the region.

#### Objectives

The specific objectives are as follows:

1. Identify mechanisms promoting and interrupting the intergenerational transmission of violence for boys/men, girls/women, and other genders
2. Explore the effect of multiple types of exposures to violence on young people’s mental health, violence experience, and perpetration
3. Examine if and how structural factors, namely poverty, poor service access and delivery, and HIV burden, contribute to violence transmission across generations
4. Investigate if government social protection policies such as child grants, free schooling or service access and provision have the potential to reduce violence transmission

### Setting

South Africans experience a high burden of all types of interpersonal violence [35–38]. In addition, vulnerable communities have been strongly adversely affected by the HIV/AIDS epidemic and high levels of poverty and inequality [39] which have been further fuelled by the COVID pandemic [40] and are established risk factors for violence and crime [41]. Furthermore, South Africa has the longest-running government social grant system in Southern Africa, allowing for long-term testing of the system’s effect on violence transmission.

### Methods and Analysis

#### Design

The study draws on data collected in 2010/11 and 2011/12 from participants aged 10–17 years old (57% female) during the Young Carers Study [42], and will follow-up participants in 2022–2023, and also recruit their oldest child (if aged 6+) and their former primary caregiver establishing a three-generational longitudinal sample.

### Mixed-methods design

The study will employ a concurrent mixed methods design [43,44] that collects, analyses and integrates both qualitative and quantitative data. Quantitative data collection will commence first, and qualitative data collection will be conducted with a subsample of the survey participants alongside the quantitative data collection.

In addition to the quantitative surveys, qualitative in-depth interviews with 30 purposely selected young people who participated in the survey (n=15 childhood violence-exposed, n=15 childhood violence-non-exposed, some HIV+ and some with poor mental health), their children (n=30) and former primary caregivers (n=30) will be conducted. Fifteen of these families will be followed up with an additional interview.

Wave three of the study is currently funded, and there are plans for future waves which are yet to be funded. Please see Figure 2 for the study design.

**Figure 2:**
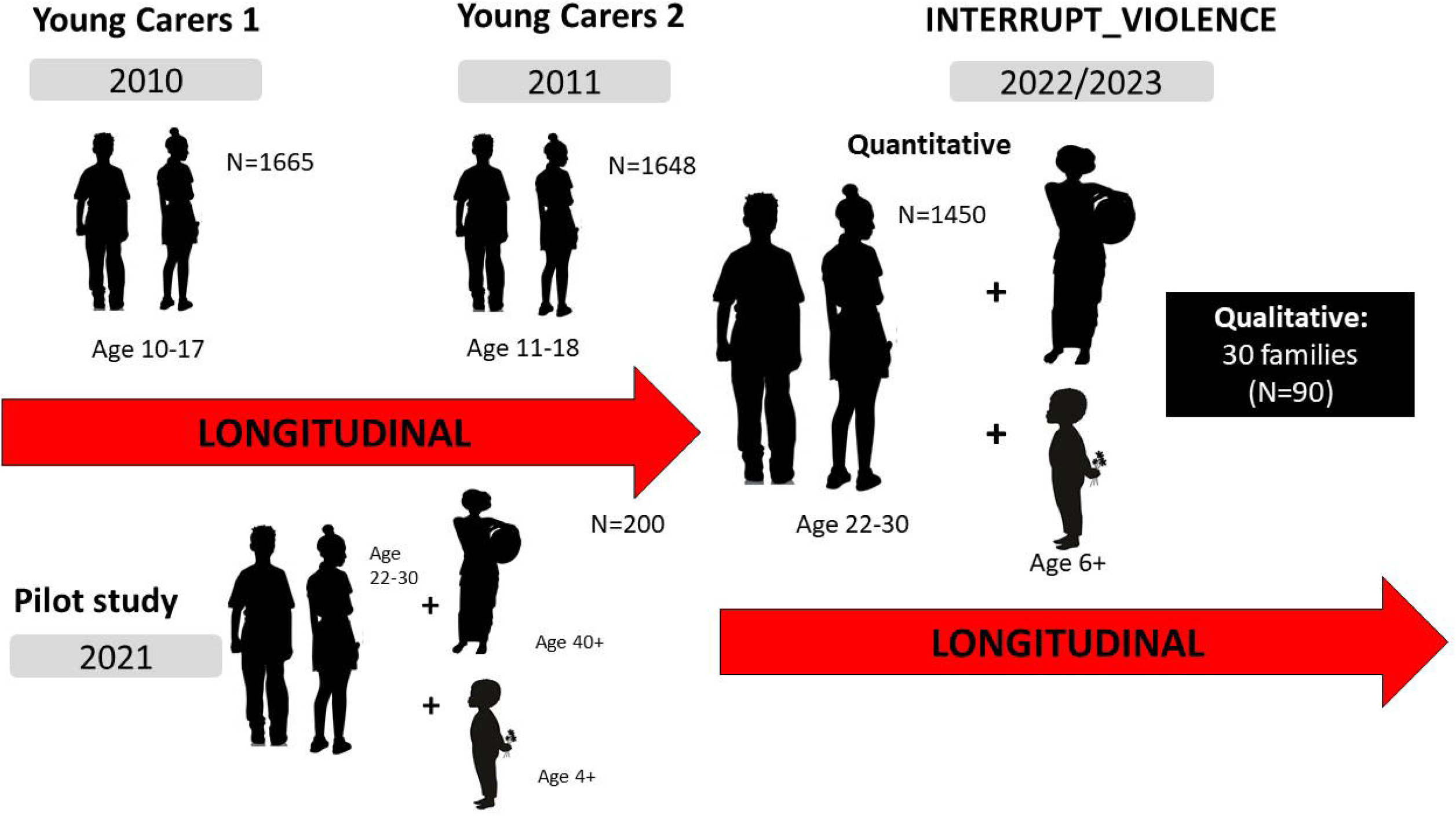
Study design of INTERRUPT_VIOLENCE.

### Quantitative Sample

#### Existing data (Wave 1 and 2)

1665 participants were originally recruited into the cohort study ‘Young Carers’ in Mpumalanga, which investigated the effects of violence in adolescence and HIV on families. Wave 1 collection was conducted in 2010/2011 with a follow-up in 2011/2012, with interviews one year apart for each participant. Adolescents were recruited from randomly selected census enumeration areas within two health districts in Mpumalanga province with antenatal HIV prevalence ≥30%. Door-to-door sampling was used to identify all households with adolescents aged 10-17. The study had good uptake with 97.5% consenting to participate and only 18 adolescents lost to follow-up at wave 2. Written caregiver consent and verbal child assent were sought for all participants. Adolescents were interviewed by local interviewers trained in working with vulnerable, traumatised, and highly stigmatised children. A 60-minute questionnaire containing validated scales measuring violence experience, risk behaviours, poverty, family illness and death, caring responsibilities, health, mental health, educational attainment, family relationships, and receipt of social protection (i.e. government grants, free schooling, school feeding, teacher support, social services receipt) was filled in by adolescents with the help of interviewers (Supplement 1). Interviews took place in spaces which ensured confidentiality and privacy. There were no financial incentives for participation, although all children received some refreshments and a certificate for participation. At each point of data collection, participants were asked whether they would like to be visited and interviewed again in the future. Mandatory referrals (n=330) to local services with follow-up support were carried out where children disclosed risk of significant harm or asked for help. The study was approved by the University of Oxford in 2009 (SSD/CUREC2/09-52), University of Cape Town (389/2009) and received additional ethical approval from the Provincial Departments of Health and Education and the South African Department of Social Development.

At wave 1 participants had the following characteristics: mean age of 13.5 years, 56% girls and 30% lived in households where a family member was ill with AIDS. In wave 2, 56.3% reported lifetime physical abuse, 35.5% lifetime emotional abuse and 9% lifetime sexual abuse. 68.9% reported any type of lifetime victimisation, and 27.1% reported multiple abuse experiences [42]. The study showed increased risk for violence experience among children in AIDS-affected families [45] and increased risk for sexual victimisation among girls who had experienced victimisation previously [46]. The study found strong associations between household deprivation and increased risk for childhood violence experience and poor child health outcomes [47].

### Wave 3 ‘INTERRUPT_VIOLENCE’

This is a follow up of the original cohort ‘Young Carers’ Cohort in Mpumalanga 10 years after the last interview. It will recruit their oldest child (if aged 6+) and their former primary caregivers (if still alive) into the study. The resulting datasets for this study will include 1) longitudinal data on violence experience and perpetration of 1665 young people collected at three time points (2010–11, 2011–12, 2022– 23) to examine links and mechanisms between violence experience in adolescence and violence experience and perpetration in young adulthood; 2) a two-generational sample of these 1665 young people and their former primary caregivers (estimated n=540) to investigate how caregiver violence experience in childhood may affect their children’s risk for victimisation and perpetration in adulthood and their uses of harsh discipline in parenthood, and 3) a three-generational sample of young people, their caregivers and their oldest child aged 6+ (estimated n=211) to explore violence transmission across three generations.

### Pilot study

Prior to the main study, an in-depth mixed-methods pilot study was conducted in Mpumalanga adjacent to the fieldwork sites to ensure all protocols, procedures and research materials were appropriate for the target population. The pilot study ran from 1^st^ July – mid October 2021 and is reported in Franchino-Olsen et al., (Investigating the transmission of intergenerational violence: A pilot study from South Africa, under review). The pilot study recruited young adults aged 22–30, their primary caregiver, and oldest child aged 4+ to test the duration of questionnaires, examine the suitability of measures for all participants, particularly children, investigated comprehension of study materials and measures for children aged 4–7 using in-depth qualitative interviews, a questionnaire and cognitive testing. Distress, recruitment, and COVID-19 protocols were also tested.

The study demonstrated the feasibility of including young children in violence research and conducting multi-generational research on violence (Woollett et al., *‘Children are like vuvuzelas always ready to blow’*: Exploring young children’s participation in violence research, under review). Children of all ages were valid reporters of their experiences of violence but children who were more developmentally and verbally advanced provided richer qualitative narratives and were able to focus for longer periods of time during the interviews. For the main study, a decision was made to interview children aged 6+. Cognitive interviews highlighted few problems with measures which were adapted or replaced in consultation with local fieldworkers; protocols were robust and worked well (Franchino-Olsen et al., Investigating the transmission of intergenerational violence: A pilot study from South Africa, under review).

### Recruitment

To re-recruit former participants, study teams will re-introduce the proposed research to local government (Ward Councillors) and community leaders (Indunas) and seek permission to return to the communities to conduct household surveys. The study will be introduced as a follow-up to the previous Young Carers Study to investigate the relationship dynamics, health, and well-being of the former participants, their former primary caregivers, and their children. Participants who previously consented to follow-up will be found using the contact details collected in 2012, attending community meetings to inform people about the return of the study and making use of neighbourhood networks and community guides to establish the whereabouts of former participants. Participants will be screened for personal information collected at wave 1 and wave 2 (e.g., national ID numbers, birth dates, names of primary caregivers, and names of primary and secondary school) to ensure they are the same participants. Once former participants have been contacted and their identity confirmed, they will be informed of the purpose of the study and have the opportunity to ask questions before being invited to consent to participation, and consent to use of previously collected data and transfer of this data from Oxford to Edinburgh in line with the South African Protection of Personal Information Act (POPIA).

In 2022, the start year of data collection, participants will be 22–27 years old. Based on low mobility of the target population, it is estimated this study will re-recruit 1600 young adults, 540 former primary caregivers, and at least 211 children, with at least 540 families in the two-generational dataset and 150 families in the three-generational dataset. In total, 2351 quantitative participants are expected to participate. This study is a collaboration between the University of Edinburgh, the University of the Witwatersrand, North-West University, and the Mpumalanga provincial Department of Social Development.

### Quantitative data collection

Original participants (young adults) who consent will be interviewed and screened for eligible children and then asked for consent for their oldest child to participate and their former primary caregiver to be contacted for study enrolment. Any child who is aged 6+ and for whom the young adult is a primary caregiver is eligible to participate in the study. Children will then be visited by an interviewer, introduced to the child survey, and given an opportunity to ask questions before being invited to provide assent.

Primary caregivers, if still alive, will be contacted by the young adult to seek permission to share their contact information with the interviewer before being visited by an interviewer, introduced to the caregiver survey, and invited to consent to participate. Data will be collected on tablets using Open Data Kit (ODK) software in the home language of the participants (English, XiTsonga or SiSwati). Adult questionnaires are approximately 180 minutes in length, child questionnaires will have a duration of 60 minutes including a 30-minute drawing task. Questionnaires include repeat measures asked at wave 1 and wave 2 follow-up to ensure availability of longitudinal data on crucial risks and outcomes that help answer the research aims. Interviewers will administer the survey. Adult participants will be able to complete sections on sensitive topics such as violence victimisation and perpetration using audio-assisted mobile interviewing.

Pre-programmed skip patterns will be used to ensure to only ask more detailed questions to participants for whom these questions are relevant. At the end of the interview, all participants will be asked for permission to be contacted again for potential future interviews following the same consent procedure. Interviews will be conducted in private in a setting agreed upon previously such as a school, a secluded bit of garden, a church, a local library. Particular care will be taken that interviews cannot be overheard or too closely observed by others. Child interviews will be conducted in pairs where one interviewer conducts the interview and the second observes to ensure privacy and safety of child and interviewer.

Where an original (young adult) participant, an eligible child, or caregiver has died, a close relative will be asked to provide information on cause of death via a verbal autopsy and a form will be completed on ODK.

### Power calculations

The sample sizes of the proposed study are predetermined by the original wave 1 study population. The maximum number of adolescents are n=1665 with an estimated n=540 of caregivers and n=211 of children based on mortality and fertility data in the area (Shoko 2016). These necessarily limit the maximum achievable statistical power. Given the theoretical complexity due to individual, family, and community factors in determining outcomes of intergenerational transmissions of violence, it would be desirable that proposed analyses should be able to detect changes for regression models with at least 20 parameters. With the longitudinal nature of the study, attrition is to be expected. Currently no evidence exists from similar settings or for similar populations that could give indications for the expected effect sizes. A recent analysis on men’s childhood violence experiences was used to identify effect sizes of predictors for perpetration in adulthood [27]. Since the number of reported effects were in the small (*r*^*2*^=.0124; Cohen’s *f*^2^= 0.01 for ‘witnessed mother abuse’) to mid-range (*r*^*2*^= .1246; Cohen’s *f*^2^= 0.14 for ‘overall effect across all childhood violence outcomes’), they provide a conservative basis for the calculations [50]. The power calculations were conducted in R (v 3.5.1) using the package ‘pwr’ [51] for linear multiple regressions for the samples of young adults, caregivers and children. Allowing as much as 25% attrition in each of the samples (n=1260 young adults/n=405 caregivers/n=158 children), this study will have at least 80% power to test models with twenty predictors or covariates to detect a moderate-range effect size of Cohen’s *f*^2^= 0.14, even in the small child sample.

### Measures for adult participants

All proposed measures have been validated for use in Southern Africa or been used extensively with similar populations in South Africa. Main outcomes are described below. For a complete list of measures please see Supplement 1 (Young Adult) and Supplement 2 (Caregiver).

#### Violence

*Child abuse* will be measured using the International Society for the Prevention of Child Abuse and Neglect’s ICAST tools for retrospective reporting of frequency of childhood violence [52] and parental reporting of frequency of past-year and lifetime exposure of their children [53]. *IPV exposure and perpetration* will be measured using the WHO domestic violence instrument which measures lifetime and past-year prevalence of different acts of physical, sexual, psychological and economic abuse, and controlling behaviour [54]. Lifetime and past-year *community violence exposure* and its frequency will be measured using items from the Social and Health Assessment (SAHA) community violence questionnaire Weissberg, Voyce, Kasprow, Arthur, & Shriver, 1991). Lifetime *peer violence victimisation and perpetration* and its frequency will be measured using the 12-item Zurich Brief Bullying Scales [55]. *Non-partner sexual violence* and *community violence perpetration* against males and females will be measured using 4 items from the UN Multi-country Study on men and violence respectively [27].

#### Health

*HIV status* will be measured using an oral swab test. For participants not wishing to test, a Verbal Autopsy Questionnaire, which is a checklist of AIDS-defining illnesses and AIDS-non-specific illnesses, will be employed [56].

#### Poverty

*Food insecurity* will be measured using the 3-item household hunger scale [57]. One additional item was added to assess if children went hungry in the household in the past 4 weeks. *Household poverty* will be measured using an index of access to the fifteen highest socially perceived necessities for households in South Africa such as soap to wash [58].

#### Social protection

*Social protection* will be measured as receipt of any of the following: government social grants, free schooling, free school feeding, free school books, free early childhood education, or access to social/health/criminal justice services following violence exposure or perpetration, being breastfeed as a child, ID/ birth certificate, mobile health messaging, bursary for studies, home-based carer, water, electricity, solar power, a community safe space, or school/community food garden.

#### Mental health

*Suicidality* will be assessed using the 5 item National Institute for Mental Health’s Ask Suicide Screening Questions [59]. *Depression* will be measured using the Patient Health Questionnaire 9-item self-report scale (PHQ-9) [60]. *Anxiety* will be measured using the Generalized Anxiety Disorder Screener (GAD-7) [61] for adults. *Post-traumatic stress and trauma exposure* will be measured using the PTSD-8 [62].

### Measures for child participants

Measures were either previously used with young child participants in South Africa and Zambia as part of the Child Community Care Study or were validated with older children and adapted for use with younger children through rewording, use of tactile aids (e.g., an empty, half-full and fully filled container with soup mix to facilitate understanding of response options like ‘never’, ‘sometimes’, ‘always’), arts-based activities with play-doh and drawings and visual aids (e.g., Figure 3). For a complete list of child measures please see Supplement 3.

**Figure 3:**
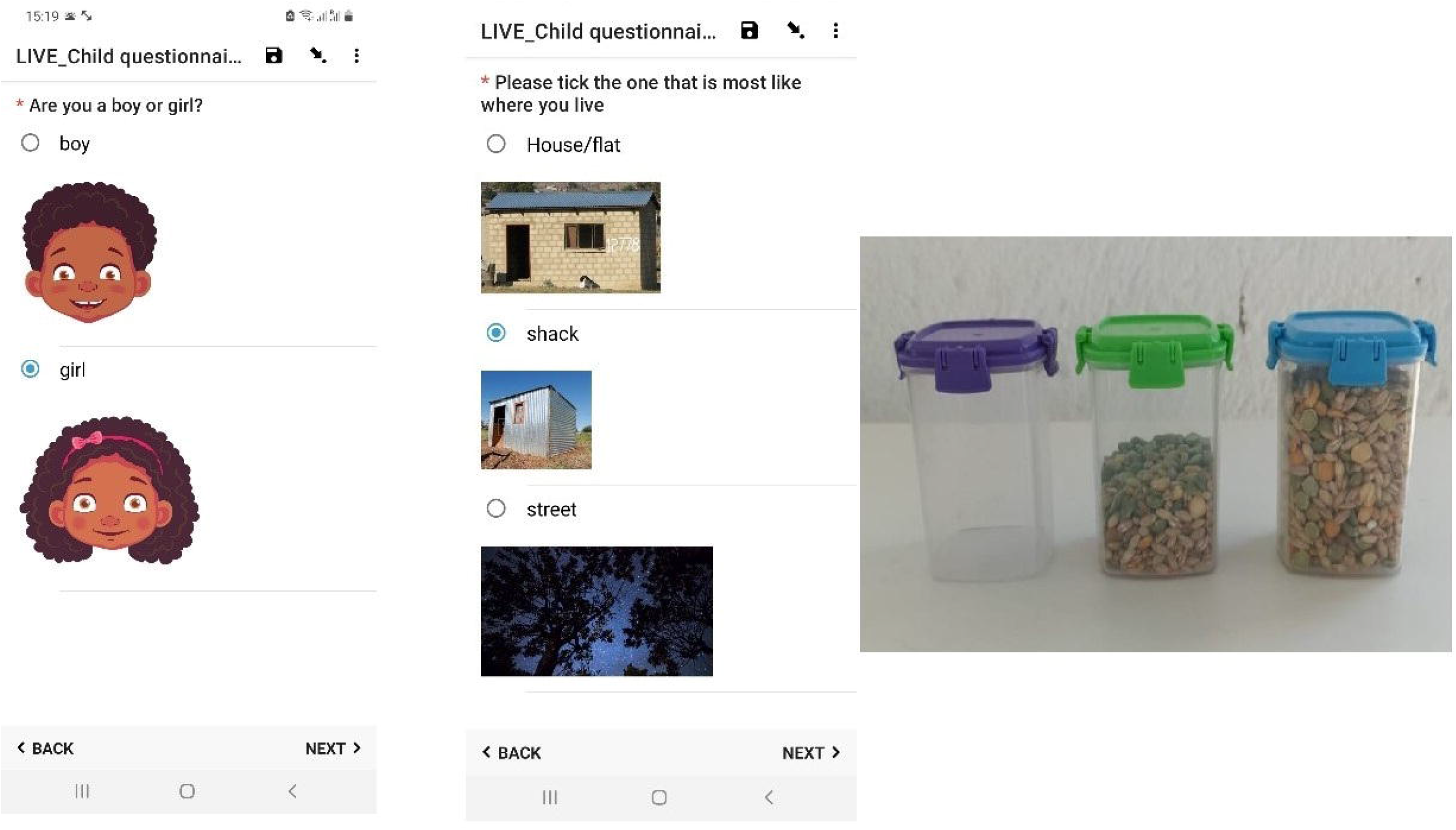
Visual and tactile aids for child questionnaires.

#### Violence

*Child abuse* and *exposure to IPV* will be measured using a six-item adapted version of the Parent-Child Conflict Tactics Scale suitable for young children [63]. *Safe and unsafe spaces and people* in the home will be mapped using the picture-based House and Community Plan which involves drawing the child’s home and using play-doh figures as those inhabiting the home [64]. Items from the Child Community Care Study will be used measuring *feelings of belonging*, treatment equal to other children in the home, and whether the child has recently changed caregiver. Children’s *attitudes and beliefs* about the acceptability of family violence are measured using 7 items from the Attitudes About Family Violence (<u>AAFV</u>) scale [65]. *Bullying* by peers and siblings will be measured with the standardized 9-item ‘Social and Health Assessment Peer Victimization Scale’ [66].

#### Support

*Positive and involved parenting* will be measured using an adapted version of the Alabama Parenting Questionnaire for young children [67].

#### Mental health

*Depression* will be measured using the 13-item Child Depression Inventory [68]. *Anxiety* will be measured using the 16-item Revised Children’s Manifest Anxiety Scale [69]. *Post-traumatic stress* will be measured using 10 items from the Trauma Symptom Checklist for Young Children [70]. *Child behaviour* including conduct problems, emotional symptoms, prosocial behaviour and hyperactivity will be measured using a 5-item version of the Strength and Difficulties Questionnaire for young children [71]. *Suicide ideation* will be measured using the 6-item MINI International Psychiatric Interview for Children and Adolescents suicidality and self-harm subscale [72]. *Resilience* will be measured using the 11-item Child and Youth Resilience Measure [73].

#### Community level factors

*Safe spaces and support* will be measured using 16 items from the Child Community Cohort Study [74]. Community violence will be assessed using 6-items from the Things I have seen and heard measure assessing witnessing of stabbings, beatings, shooting, sexual assault and having something stolen [75].

### Qualitative interviews

Qualitative in-depth interviews will be conducted with 30 young adults who participated in the survey (n=15 childhood violence-exposed, n=15 childhood violence-non-exposed), their children (n=30) and former primary caregivers (n=30) who have SiSwati as a first language. These will be supported by a semi-structured guide to explore participant’s experience/perception of violence and views/beliefs regarding the intergenerational transmission of violence. Interviews will cover content such as parenting styles, relationships and attachment to one’s caregivers, exposure to violence in the home as well as the community, HIV, poverty, means of coping with violence, resilience, mental health, and access to services. In-depth interviews will be conducted face-to-face, in a private space, at a convenient time for participants.

In order to facilitate these discussions, a range of arts-based approaches will be used. These will include drawings: squiggle drawing, kinetic family drawings (KFD) [76], road of life [77] and sandboxing [78]. Drawing is thought to help participants feel more relaxed with the interviewer, organise narratives and promote expression [79,80]. More information on the arts-based methods can be found in Woollett et al., (“Those drawings have messages that the children wouldn’t be able to tell you straight” - using arts-based approaches in research on violence, under review).

Research that utilises non-verbal methods, such as drawings and sandboxes, is increasingly recognised as particularly ethical as it offers research subjects active participation in the research process, authenticating their voice through their engagement. It offers more developmentally appropriate means of accessing data, diminishing stress in the child/adolescent-adult interaction and provides a more comfortable method of engagement than language [82–85]. Non-verbal methods thus offer participants an easier platform for communication, especially about sensitive and personal issues such as violence.

### Data storage

All research data will be collected on tablets and stored in a secure environment at the University of Edinburgh. Daily checks will be carried out on consent and survey forms against the sampling list. Surveys are programmed to avoid accidental skips which result in missing data. Daily quality checks will be carried out on all questionnaires to detect inconsistencies and inaccuracies. Qualitative and quantitative data will be pseudonymized using unique barcodes for each participant and family. Identifying information will also be collected and stored encrypted and separately to allow linkage of records at the end of the study and to facilitate referrals.

Referrals for distress and mandatory reporting are based on an algorithm programmed into the survey questionnaire and through interviewers based on referral criteria. These are checked daily to ensure urgent referrals are contacted immediately.

### Quantitative data analysis

All quantitative analyses will be carried out in STATA 17.1 and Mplus 8.8. For the longitudinal and multi-generational samples, logistic regression analyses, path analyses, and structural equation models are planned. Latent growth curve modelling will be considered as an approach to investigate changes in violence experience over time for the three-wave longitudinal study of young adults. Propensity score matching or difference in difference estimation will be considered to investigate the effects of social protection on violence transmission. Missing data will be an important issue in this proposed longitudinal study. Full maximum likelihood estimation will be used to address all missing data as this is currently assumed to be the most rigorous method for missingness [86]. Where missing data are minimal and only on individual items, imputation methods may be considered if appropriate.

### Qualitative data analysis

Interviews will be translated from SiSwati to English and transcribed by the qualitative interview team. An initial (top-down) broad coding framework will be developed based on the study research questions using a thematic approach [87]. Using MAXQDA 2021, broad codes will be applied to all transcripts by two researchers, then the team will establish a system of fine codes that emerge inductively from the data (bottom-up), deriving meaning from the data itself rather than imposing pre-formed ideas [88]. To ensure intercoder agreement, fine codes will be developed by printing out a full set of excerpts (from each data set) related to each code for each transcript and identifying sub-themes emerging from the data. All transcripts will be double coded. An analytical summary will be developed on each study objective using illustrative quotes to support key emerging conclusions. The findings and analytical reports will be critiqued by the group to guarantee research findings; highlighting the reality of the transcripts rather than simply one researcher’s view of the data. Structured reflection notes will serve as an additional decision-making “audit trail” throughout the process. Qualitative biases will be addressed by this team approach to data analysis, accounting for inter-coder reliability rates. Analysis of drawings and sandboxes, will be assisted via facilitated focus group discussions.

### Community and public involvement

The study will be supported by two community advisory boards (CAB)—one for each study site—consisting of adolescents aged 10–17 and adults. CAB members will be drawn from the communities in which the research takes place through recommendations from leaders, study participants, and research staff. The CABs serve to provide community input into the research and bring specific, unique expertise to the research process by informing study teams of local issues or concerns that can affect the conduct and successful implementation of the research and will advise on issues of particular concern to children and adolescents. The CABs will inform the research team about information and misinformation pertaining to the study in the community and disseminate authorized study information. The research team will use the RIGHTS framework to engage the CABs (Oliveras et al, 2018).

### Ethics and dissemination

Ethical approval was granted by the University of Edinburgh School of Social and Political Sciences (No: 264227), University of the Witwatersrand Human Research Ethics Committee (M190949), North-West University Health Research Ethics Committee (NWU-00329-20-S1), and the Provincial Department of Health Mpumalanga (MP-202012-003).

In order to minimise harm to particpants, the study will not be presented as a study about violence or HIV but instead as a study about the well-being of young adults and their children in the context of their family and community background. Questions on perpetration and victimisation will be embedded among other survey questions related to the well-being of young adults and their children in line with WHO guidelines on violence research [89]. Participants will be advised not to discuss the questions on perpetration and victimisation with others in order to avoid stigmatisation, retribution, and misunderstanding. Interviewers will be trained in recognising distress and trauma of participants. Emotional support and empathy will be offered and breaks or postponement of interviews granted following guidelines on conducting research on violence [90].

A social worker will be employed by the research team on a full-time basis for the duration of the fieldwork. The social worker will be expected to manage mandatory referrals raised as part of the interviews or in interactions with participants commensurate with the Children’s Act (2005) [91]. They will also be expected to support families and participants who seek help e.g., around gaining birth certificates, access to grants or dealing with distress. Their main responsibility is to facilitate referrals to local specialist services, which can provide support to families and children on a long-term basis. Prior to the commencement of data collection, the social worker compiled a community profile to identify referral agencies and build rapport with service providers in the area. This ensures that they are familiar with all the resources in the community to streamline referral procedures. The social worker will receive regular supervision sessions to attend to their emotional well-being and to discuss challenges encountered as part of the work. Further detail on the study’s social worker will be described separately.

This study will be interviewing children aged 6+ which presents risks and challenges in relation to safeguarding, participant’s rights to participation [92], and their cognitive ability [93]. The research team has given considerable thought to developing procedures to ensure that interviewers are appropriately qualified to interact with children this young and that the questionnaire is suitable for the age groups described. It is the utmost priority of the research team that children are protected, and where they disclose violence or distress, are on-referred to appropriate services.

For disclosures of child abuse victimisation or perpetration (where the child can be identified), the social worker will be available to assist with containing any feelings of distress and managing referrals. Decisions about which services to contact as well as processes for referrals will be discussed with the the child (if the child has made the disclosure) in the first instance, and the non-perpetrating caregiver in the second instance. These reporting requirements are highligthed in the informed consent procedures. All interviewers will receive extensive training on dealing with disclosures of violence experience, particularly by child participants and in providing evidence-based alternatives to harsh punishment to caregivers who seek advice. It should be noted that disclosures of victimisation and perpetration are high in South African research studies despite the mandatory reporting requirements [42].

For questions about perpetration, the amount of detail asked will be minimised so that individual victims cannot be identified from the research. This is in line with best practices on researching violence perpetration [90]. Moreover, partner violence perpetration questions and non-partner violence victimisation questions will be audio- and computer-assisted, allowing for self-completion for adult participants. Any additional information provided by the participants will then be understood as a request for help. All information disclosed in the research process will remain completely confidential unless the victim is a child and is identified as such by the participant and described in a way that allows the research team to identify the child. In this case, referral procedures will come into effect and further steps will be discussed with the participant unless doing so will put the child at risk of harm. Where a referral might put the child at risk of harm, the research team will consult extensively with local services to ensure the safety of the child.

### HIV status

Participants are encouraged to disclose their HIV status in the interview if they are aware of it but are not required to do so. Further, voluntary HIV self-testing will be available for all adult participants by means of an oral HIV self-testing kit. All interviewers have qualifications in voluntary HIV testing and counselling. Protocols are in place to link participants with positive HIV test results and their children to clinical care. HIV treatment and care are free of charge in South Africa for all people living with HIV [94].

### Reimbursement

All adult participants will receive a reimbursement for their time in the form of a ZAR50.00 gift voucher and refreshments. Child participants will receive a pack consisting of stickers and some refreshments.

### Training of fieldworkers

Criminal background checks and reference checks will be carried out on all interviewers prior to taking up employment. Intensive training will be provided to all interviewers for five weeks and will include sessions on 1) gender norms, intimate partner violence, sexual consent, and intersections between violence, HIV and mental health, 2) values clarification around gender, violence and children, 3) children’s rights and violence against children, 4) child development, 5) interviewing techniques, 6) study protocols, 7) making children feel comfortable and building rapport, 8) non-verbal cues and body language, 9) HIV testing and counselling, and 10) implementation of containment skills where participants show strong emotional responses. Training will follow an experiential and authentic learning approach [95]. First, skills will be mastered during class simulations, followed by practicing those skills in a real world setting with a low risk population. We also expect to provide frequent refresher sessions throughout the duration of the fieldwork based on gaps or protocol inconsistencies observed in field and through regular focus group discussions with the fieldwork team.

Qualitative interviewers will receive a 40-hour intensive experiential training on the study methods and implementation of multigenerational violence research, including interviewing young children within families. The team will be closely supervised and mentored in the use of arts-based methods. Any misconduct or pressuring of participants will be grounds for dismissal. More information on the training of interviewers can be found in (TRAINING PAPER).

Results will be shared with local government, international organisations and the public through diverse engagement activities including social media, radio, presentations and the community advisory boards. Results will be published in peer-reviewed journals, at scientific meetings, and policy briefs. All publications will be made available on the project website. Anonymised datasets will be made available 10 years after the end of the project through the UK Data Service.

### Limitations

Despite its strengths, INTERRUPT_Violence has limitations. The main risk to the study is around attrition of participants. Levels of attrition are difficult to predict and will impact overall sample size. Second, there is a risk of research fatigue, particularly for participants who participate in both the quantitative and qualitative research, which may be mitigated through participatory arts-based methodologies in the qualitative component. Third, it is possible that the involvement of a study social worker and the mandatory referring of participants to other services will impact findings. However, previous research has shown negligible impact of these interventions [96] and only a small number of our participants will receive more than one counselling session. Recording those who are referred and those who receive long-term intervention will allow investigation of the impact of these service provisions on outcomes.

## Conclusions

Violence has major impacts on society and health across the life course and generations. The global agenda of the United Nations has finally made the eradication of violence one of their key strategy targets as part of the Sustainable Development Goals. Longitudinal research on the underlying mechanisms of the transmission of violence across generations is urgently needed to prevent and respond to violence and to develop effective, evidence-based programmes and policies, particularly for sub-Saharan Africa. The study will recruit a three-generational longitudinal sample of vulnerable families in a Southern African context and will be the first of its kind to focus on intergenerational violence transmission. It will also be the first study to develop an empirically generated theoretical framework using robust methodology and to examine government social protection policies for violence prevention. This study should have broad applicability to violence research and prevention efforts in many countries. The proposed research will therefore address multiple urgent research gaps and advance knowledge to address this important public health concern through ground-breaking scientific methods.

## Data Availability

Anonymised datasets will be made available following the end of the study through the UK Data Archives.

## Acknowledgements

Interrupt_Violence Advisory Committee: Elsinah Mhlongo, Alexander Butchart, Karen Devries, Heidi Stöckl, Lorraine Sherr, Mark Boyes, Andrea Gonzales, Lucie Cluver and Michael P. Dunne.

## Author statement

FM designed the Interrupt_Violence study with substantive input from NC, NW, HFO, MS, CT, AF and KM. FM drafted this manuscript. All authors read and approved the final manuscript.

## Ethics approval and consent to participate

The INTERRUPT_VIOLENCE study has been approved by the University of Edinburgh School of Social and Political Science Research Ethics Committee (264227), the University of the Witwatersrand Human Research Ethics Committee (M190949) and North-West University Health Research Ethics Committee (NWU-00329-20-A1). Further ethical approval was granted from the Mpumalanga Department of Health (MP-202012-003). All adult participants in this study provide informed written consent prior to involvement in the study. For child participants, primary caregivers are provided with information about the study and consent to their child’s participation. Children provide written assent prior to involvement in the study. Consent and assent procedures were approved by all involved Ethics Committees.

## Footnotes

1 https://sustainabledevelopment.un.org/sdgs

