## Supplement 1 for "Interrupting the intergenerational cycle of violence: protocol for a three-generational longitudinal mixed-methods study in South Africa (INTERRUPT_VIOLENCE)"

Supplement 1: Young Carers 3 – Young adult questionnaire

| | Used in 2010/11 and 2011/12 | INTERRUPT_VIOLENCE 2022-2023 |
|---|---|---|
| **ABOUT YOU** | | |
| Demographic Information | Child age, gender and basic demographic information were asked using items from the South African Census 2001 [1]. Household structure was measured using a Household map, devised for complex extended family structures and used in our previous studies with AIDS-orphaned children and young [2]. This uses a picture-based tool to determine household members, relationship to children, age, gender, and living arrangements (i.e. who sleeps in which rooms). It also determines whether households have access to kitchen, bathroom and toilet facilities. Items from the South African Census 2001 determine whether household water supply is from tap, community tap or other source. This was suggested by the Departments of Health and Education, and may impact on experience of caring for Young Carers. | Age, home language and basic demographic will be asked using four items [3], South Africa General Household Survey [4], and the South Africa Demographic and Health Survey [5]. Questions on sexuality, gender and sex assigned at birth were modelled on the US Consortium of Higher Education LGBT Resource Professionals Self-Study [6]. |
| Education | Education items were developed with the South African Department of Education. Some items from the 'Young Lives' study [7] were used to determine some education outcomes, others were identified by NACCA and NGOs. Items included: age of school enrolment, grade for age, repetition of grades and reasons for repetition, failure of grades and reasons for failure, school dropout and reasons. We measured number of different schools attended, and migration within and between provinces. We also examined ability to pay school fees. For young carers, we measured daily allocation of time for school and homework. Items also examined missing school due to responsibilities. Further items examined whether children felt that their teacher understood their home | Six items will assess educational background. Highest level of education attained will be measured using items from the IMAGES Study South Africa 2018 [8].<br><br>Additional items will focus on completed schooling: school dropout and reasons for drop out, school interruptions.<br><br>Additional items will focus on tertiary education where applicable: highest further education; drop out and reasons for drop out, and they are/were receiving scholarship.<br><br>Violence exposure at school was measured with one item. |

| | | |
|---|---|---|
| | situation, and whether children have accessed counselling at school. | |
| Legal Status | | 5 items will ask about citizenship status if they have a birth certificate, ID book or passport and the reasons for not having an ID or birth certificate. |
| **YOUR HOME** | | |
| Your Home | Poverty was measured using a range of tools. Items from the South African Census (Stats SA) determined whether children lived in formal, informal (i.e. shacks) or traditional structures. Poverty was also measured by measuring access to the top 8 socially-perceived necessities for children, as identified by the Centre for South African Social Policy in the 'Indicators of poverty and social exclusion project' [9], and endorsed by over 80% of the South African population in a nationally-representative survey (the South African Social Attitudes Survey 2006) [10]. These include items such as 'enough clothes to keep you warm and dry' and '3 meals a day'. | 21 items will be used to identify size of the home, type of home, number of people living in it and the relationships to the participants based on items from the South African Census [3] and IMAGES Study [8]. These cover if house is owned or rented, number of rooms, kitchen, toilets, number of chronically ill people in the home, number of employed people, whether adults and children share beds, the household water and electricity source and how many days either were not available in the past month. |
| Employment | Household employment was measured in the Household map, where employment of any adult or child in the household was identified, and distinguished regular, part-time and seasonal/irregular work. | Six items from the IMAGES Study South Africa [11] assess employment and household income focusing on employment of respondent, whether the work is seasonal, part-time or full-time, the monthly household income, number of adults and children dependent on the household income. |
| Making-ends-meet | Food insecurity was measured using 2 items from the South African National Food Consumption Survey (1999) [12]. | Food insecurity will be measured using the three item Household Hunger Scale (HHS). This is a subscale of the HFIAS (Household Food Insecurity Scale). It consists of 3 items and has demonstrated the potential for cross-cultural validity. The HFIAS has been validated throughout different countries, including South Africa [13]. One additional item is included to assess whether children had enough food in the past 4 weeks. |

| | | |
|---|---|---|
| | | 8 items from necessities approach of Barnes and Wright [14] and 6 additional items will measure what necessities the household was able to afford in the past month. These will include three meals a day, sanitary products, household cleaning products, electricity, water, shoes amongst others.<br><br>4-items from the Sinovuyo Teen Study will assess a household's financial management including membership in e.g. a stokvel or burial society, borrowing money and ability to cope with financial shocks [15]. |
| Government Grants | Receipt within the household of the major forms of *social security transfers* (Child Support Grant, Foster Care Grant, Pension, Disability Grant and Care Dependency Grant) was measured, and reasons for *non-receipt of grants* assessed using the most-identified causes of non-receipt in a study of welfare access in South Africa [16] (5 items). Assistance in the forms of *food parcels* and *soup kitchens* was assessed using items devised by the National Action Committee for Children Affected by AIDS (NACCA), and followed by items to assess reliability of these services (6 items). Respondents identified whether their *school* was a no-fees school, or whether they had a fees exemption (3 items). They also identified whether they had state-provided school uniform, school transport, free school text books or school feeding scheme (items identified by the Department of Social Development). (3 items). Further items measured access to *social worker* and frequency of visits. *Home-Based Care* services were measured using items to determine frequency of visits and services provided to the child. Children also reported access to an ID book, birth certificate, and death | Receipt within the household of the major forms of social security transfers (Child Support Grant, Foster Child Grant, Care Dependency Grant, Disability Grant, Grant in Aid, government housing subsidy, Old Age Pension) will be measured. An additional item will ask if the household received the special COVID-19 Social Relief of Distress grant and the one time R700 grant provided in 2020 as a result of the COVID-19 pandemic. |

| | | |
|---|---|---|
| | certificate of parents in case of grant requirements (items suggested by DSD) (3 items). | |
| Communication | | Two items will assess whether households have access to a mobile phone or smart phone, TV, radio and computer/internet and whether and what type of health messaging they may have received through their devices in the past year. |
| **YOUR FAMILY AND FRIENDS** | | |
| Social Support | Social Support was assessed using the standardised Social Support Scale [17], measuring social support for urban adolescents in each microsystem of family, peers and school. Psychometric properties were acceptable: α=.63 in a US study. This scale has been used in Cape Town [18] and in our prior study of AIDS-orphaned children. The scale was adapted to replace 'mother' and 'father' with 'caregiver'. (24 items) and showed an α.76 in our prior study. | The 8-item Modified Medical Outcome Study Social Support Survey (mMOS-SS) will be used to measure social support [19] which has similarly excellent validity compared to the original 18-item MOS-SS. The MOS-SS was designed to measure social support received by persons with chronic conditions. It has good internal consistency α=0.91to 0.97 and test-retest reliability α=0.72 to 0.78 [20]. It has been used in our previous studies in South Africa with good success [21,22]. |
| Support from churches and traditional healers | Family contact with traditional healers was measured with three devised items. Social support through the church or religious leaders was also included in the social support measure. | Not retained |
| Support from School | Support gained from schools included a measure of social support in the Social Support Scale. | Not retained |
| **YOUR CHILDREN** | | |

| | | |
|---|---|---|
| Presence of children and involvement with children | NOT MEASURED IN EARLIER WAVES | For those assigned male at birth, three items will assess whether they have been told by a woman that they made her pregnant and how many of these children they accept they are the father of, and about their involvement in bringing up these children.<br><br>For all participants, four items assess how many children they care for like a parent, their relationship to the oldest child they are caring for like a parent (index child), the name of this child and their age. One item assesses which roles they provide in the index child's life ranging from providing money for the child to helping with daily tasks and disciplining them.<br><br>Seven items assess the number of biological children not living with the participant, their involvement with these children, how much support they provide.<br><br>One item will assess whether they treat all children in their household equally<br><br>Four items assess the index's child's gender, how easy they are to care for, how they feel about the index child and how much time they spend playing with the index child. |
| Child mortality | NOT MEASURED IN EARLIER WAVES | Four items will assess if any of their children have died, how many, how old they were when they died and the cause of death. |
| Pregnancies | NOT MEASURED IN EARLIER WAVES | Eight items for women assess the number of times they have been pregnant, how many live births, miscarriages and terminations they had, age of first pregnancy, access to MomConnect and antenatal care and whether the pregnancy was planned. |
| Index child protection | NOT MEASURED IN EARLIER WAVES | Two items will assess whether the index child was exclusively breastfed for 6 months, and whether their birth was registered |
| Fathering | NOT MEASURED IN EARLIER WAVES | For men, two items will assess whether they attended antenatal care appointments with their partner and whether the pregnancy had been planned. |

| | | |
|---|---|---|
| Children's Education | NOT MEASURED IN EARLIER WAVES | Nine items will assess whether the index child attends crèche or school, whether they receive a fee exemption, receive free school books, school meals, school trips, school uniform or school transport. |
| Parental Stress | NOT MEASURED IN EARLIER WAVES | Parenting stress will be assessed using the 18-item Parenting Stress Scale (PSS) which was developed as an alternative to the 101-item Parenting Stress Index [23]. It provides a measure that considers positive aspects of parenting as well as the negative, focusing on traditional "stressful" aspects. The PSS has previously been used in a longitudinal cohort study in South Africa [24]. |
| Attitudes to discipline | NOT MEASURED IN EARLIER WAVES | Three self-developed items will assess attitudes towards disciplining children. These cover endorsement of physical discipline, gendered discipline and parenting similarly vs differently to one's own parents. |
| Early Childhood Parenting | NOT MEASURED IN EARLIER WAVES | 38 items from the Comprehensive Early Childhood Parenting Questionnaire (CECPAQ) support, structure and stimulation sub-scales will be used to assess engagement with the index child, creating play opportunities for the index child, disciplining the child and consistency of parenting behaviours [25]. This measure has not been previously used in South Africa but was piloted extensively with some cultural adaptations e.g. toys and games specific to South Africa.<br><br>7 items from Involved Parenting subscale of the Alabama Parenting Questionnaire [26] measured parental involvement in the index child's activity. These have been previously used in South Africa with good validity and reliability [21]. |
| Child Abuse and Neglect (Disciplining and protecting your children) | NOT MEASURED IN EARLIER WAVES | 39 items from the ISPCAN Child Abuse Screening Tool parent self-report (ICAST-P) will be used to self-report on the use of physical and emotional punishment and the child' experience of neglect, sexual abuse, domestic violence and sibling bullying. Items on positive discipline are also included [27]. |

| | | |
|---|---|---|
| Attitudes to corporal punishment | NOT MEASURED IN EARLIER WAVES | 4 items will be asked to measure their attitudes on corporal punishment and whether they think girls should be disciplined differently than boys and if they treat their children differently to how they were treated as a child. |
| The Health of Your Children | Items from the South African Demographic and Health Survey (2003) [28] the World Health Organisation Stop Tuberculosis Team, and the Kwa-Zulu Natal Department of Health TB symptom guidelines, were used to provide symptoms of pulmonary TB. Access to and adherence to TB testing and medication was measured in 4 items, based on items from the REACH (Researching Equitable Access to Healthcare) 4-province study [29].<br><br>The Health Systems Trust South African Health Review [30] was used to identify common diseases of childhood, and items from the DHS will be used to identify other illnesses (16 items total). Chronic illnesses were measured by prevalence of asthma, epilepsy and diabetes, minor ailments includes cold/flu and worms, acute conditions includes pneumonia, diarrhoea, burns and physical disability such as visual disability. 10 items from the 12-item WHO Disability Assessment Schedule 2.0 (WHODAS 2.0) will be used to assess disability and overall functioning [31]. It includes items on cognition, mobility and self-care. Each item is scored on a 5-point scale, where 0 = no difficulty and 4 = extreme difficulty. WHODAS 2.0 found to be very reliable cross culturally by test-retest studies (WHO, 2010). | Seven items covering injuries and burns, diarrhoea and vomiting, pneumonia or bronchitis, skin conditions, general body pains, cold/flu and COVID-19 will be retained from the previous questionnaire. Pulmonary TB will be measured using the TB symptom assessment used in the baseline and follow-up research. Access to health care will be assessed with 8 items from the REACH study [29].<br><br>Disability will be assessed using 9 items from the Short Set on Functioning Washington Group Questionnaire (WG-SS) [32]. These cover visual, auditory, mobility and cognitive disabilities. |
| HIV | NOT MEASURED IN EARLIER WAVES | 9 items adapted from Sinovuyo Teen Study (2015) will be asked to assess if their children have been tested for HIV, if yes, are they positive, if yes, are they taking any medication, did they need to go to hospital in the last month, did they tell their child that they are |

| | | |
|---|---|---|
| | | HIV-positive, do they know their child's CD4 and if they can tell us [33]. |
| **YOUR RELATIONSHIPS** | | |
| Your Relationships | NOT MEASURED IN EARLIER WAVES | Participants' relationship status and experiences will be measured using items from the IMAGES Study South Africa 2018 [8]. The items will assess what their main motivation in staying in the relationship if they have one, if their partners drink alcohol, use drugs or have HIV, how many one night stands they had in the past year and if any of their sexual partners were from the same sex. These items also assess the nature of the relationships ranging from main partners to casual partners and side partners. |
| Gender Equitable Norms | NOT MEASURED IN EARLIER WAVES | The Gender Equitable Men (GEM) Scale measures attitudes toward gender norms in intimate relationships or differing social expectations for men and women [34]. 8 items from the Inequitable scale will be used and 2 additional items specific to the South African context on lobola and rape were added. Each item is scored on a 3-point scale, where 1 = agree, 2 = partially agree, and 3 = do not agree. The estimate of internal consistency = .81, but it is expected to vary according to the setting. This measures has been previously used in South Africa [35,36]. |
| Relationship Control | NOT MEASURED IN EARLIER WAVES | The Sexual Relationship Power Scale (SRPS) measures power within sexual relationships for both men and women. 8 items of the Relationship Control subscale are used in the questionnaire. Items are scored on a 4-point Likert scale, with 1 = strongly agree, 2 = agree, 3 = disagree, and 4 = strongly disagree) [37]. |
| Intimate partner violence | NOT MEASURED IN EARLIER WAVES | *IPV perpetration* will be measured using the 19-item questionnaire from the UN Multi-country cross-sectional study on men and violence and Asia and the Pacific [38,39]. Questions will ask if the behaviour happened in the past 12 months, if yes, the frequency in the last year and the frequency in the woman's lifetime.<br><br>*IPV experience* will be measured using the 25 item WHO domestic violence instrument from the WHO Multi-Country Study on |

| | | |
|---|---|---|
| | | Violence Against Women [40] for past year and lifetime exposure. Five additional items assess IPV experience in pregnancy. For women, additional questions cover whether they have sought help, whom they have sought help from and what the response to their disclosure was. |
| Community violence perpetration | Not measured in earlier waves | 8 items from WHO community violence perpetration instrument developed by Tremblay et al., [41] and adapted by Jewkes & Morrell [42] will be used to assess whether a participant has taken part in violent activities within the community in which they live . |
| Risky Sex | Items from the National survey of HIV and risk behaviour amongst young South Africans [43] were used to measure sexual activity (screening item), HIV knowledge, self-perceived risk of HIV-infection, self-efficacy/agency in relation to condom use, age of sexual debut, concurrent sexual partners, frequency of condom use, transactional sex, sexual activity under the influence of alcohol and under the influence of drugs, and self-perceived risk of HIV. Items from this survey were also used to measure forced sex (by verbal and physical force) and pregnancy (14 items).<br><br>Items from the South African Demographic and Health Survey (2003) [28] were used to measure sex with older partners and use of contraception. (2 items) | Items from the National survey of HIV and risk behaviour amongst young South Africans [43] will be retained to measure sexual activity (screening item), age of sexual debut, frequency of condom use, sex with an older partner, sexual activity under the influence of alcohol and under the influence of drugs, and self-perceived risk of HIV. Items from this survey were also used to measure forced sex (by verbal and physical force) and pregnancy (14 items).<br><br>Women's transactional sex with men will be measured using a measure from a larger study on gender-based violence and HIV infection in Soweto [44]. Men's transactional sex with women who are not their wife or main partner will be assessed using the 25 item female version adapted for men [45] . |
| Non-partner sexual violence victimisation | NOT MEASURED IN EARLIER WAVES | Non-partner sexual violence against males and females perpetrated by men will be measured using 5 items from the UN Multi-country Study on men and violence [39]. There will be follow-up questions after the items such as how old they were |

| | | |
|---|---|---|
| | | when the incident first happened, who did it, how old they were when the last happened. |
| **MY COMMUNITY** | | |
| Community Violence | Community-level trauma was measured using items from the Child Exposure to Community Violence (CECV) Checklist [46], adapted to reflect commonest community traumas in South Africa, as identified by national police statistics [47]. | Lifetime and past-year community violence exposure (witnessing, experiencing) and its frequency will be measured using 14 items from the Social and Health Assessment (SAHA; [48] Exposure to Violence Scales (Victimization by Community Violence and Witnessing Community Violence) with 5-point response options ranging from 'none' (0) to 'ten or more times' (4). The internal consistency of the scale in the US study is found to be good (α = .78 and α = .89). |
| Community Participation | NOT MEASURED IN EARLIER WAVES | Three items from the adapted Critical Consciousness Scale [49] will assess community participation with regards to how much participant agree that community members would intervene if they saw a fight, a child skipping school, or someone selling drugs on the street. |
| Community Cohesion | NOT MEASURED IN EARLIER WAVES | Six items adapted from the Stepping Stones Questionnaire [50] will assess community cohesion with a specific focus on safety e.g. walking around at night. A 4-point likert-scale will assess participant's agreement with the statement of how safe the community feels. |
| Natural disasters | NOT MEASURED IN EARLIER WAVES | Three self-developed items will assess participant's experiences of drought, flooding and fire. |
| Trauma | NOT MEASURED IN EARLIER WAVES | 10 items are included from The Harvard Trauma Questionnaire (HTQ) which is a checklist written by the Harvard Program in Refugee Trauma to assess trauma and torture [51]. Among the items, there are imprisonment, torture, unnatural death, kidnapping. |

| YOUR HEALTH | | |
|---|---|---|
| Depression Symptoms | The Child Depression Inventory (short form) (10 items) [52] was used in our previous studies of AIDS-orphanhood, and showed an acceptable α=.67 (2006) and α=.69 (2009). The CDI has strong psychometric properties, and has been used in multiple South African populations [53], including an adapted version ([54], which was validated against the Beck Depression Inventory (r=0.81). Internal consistency ranged from .71 to .94. [55] | The Patient Health Questionnaire (PHQ-9) is a nine item self-report screening tool for accessing the severity of depression based on DSM-IV criteria and scored on a 4-point scale where 0 = not at all, 1 = several days, 2 = more than half of the days and 3 = nearly every day over the last two weeks [56]. This has been previously used in South Africa [57]. |
| Disability | Two items measuring *physical disability* included visual disability and others, but not learning disabilities as these may be unreliable in self-report. | The 6-item Washington Group Short Set (WG-SS) on Functioning will be used to assess disability status [58]. This is a comprehensive measure to determine disability in six domains of functioning: seeing, hearing, walking/climbing steps, remembering/concentrating, self-care, and communication. Each item is scored on a 4-point scale where 0 = no difficulty, 1 = some difficulty, 2 = a lot of difficulty, and 3 = cannot do at all. The WG-SS has been extensively tested and validated and used in over 80 countries [58]. |
| Anxiety | The Children's Manifest Anxiety Scale – Revised reduced items (14 items) scale was used to measure anxiety [59]. In the previous orphan study, the full scale showed an α.80 (2005), and the reduced scale showed α .75 (2006) and .80 (2009). The RCMAS has been standardized in US populations, shows good internal consistency and test-retest reliability (.68 after 9 months) [60] | The General Anxiety Disorder (GAD)-7 seven item questionnaire measures symptoms of GAD following DSMV-IV symptom criteria for GAD and assigns scores of 0, 1, 2, and 3, to the response categories of 'not at all', 'several days', 'more than half the days', and 'nearly every day', respectively. Using the threshold score of 10, the GAD-7 has a sensitivity of 89% and a specificity of 82% for GAD [61]. This tool and its 2-item form have been used in South Africa [62,63] |
| Suicide Screening Questions | Mini International Psychiatric Interview for Children and Adolescents suicidality and self-harm subscale [64] (5 items) was used to measure suicide ideation. The MINI-Kid has been extensively validated in developed world populations, and shows strong internal consistency and test-retest reliability [65]. | 5 items are from the National Institute for Mental Health's Ask Suicide Screening Questions (ASQ) Toolkit [66]. It was developed as a brief, straight-forward tool to identify potential suicide risk. It includes items like 'in the past few weeks, have you wished you were dead' and 'have you ever tried to kill yourself'. |

| | | |
|---|---|---|
| Behavior | Child Behaviour Checklist [67] delinquency subscale (12 items) α=.62. The CBCL-YSR has been normed on a mixed-ethnicity US population, and was used in the Family Health Project study of orphan well-being in the USA and in South Africa [68]. Alphas ranged from .71 to .95 [69,70], and test-retest values from .47 to .79. The CBCL-YSR is commonly used as a 'gold standard' in testing convergent validity of other instruments.<br><br>Strengths and Difficulties Questionnaire [71] conduct problems subscale (5 items). The SDQ is well-validated, and has been translated into 51 languages, including isiXhosa and isiZulu [72]. In a norming study with 10,438 children [73], the SDQ showed mean α=.73 and .62 retest stability at 6 months.<br><br>Two devised items on carrying of weapons were added – these were adapted from the National Primary Schools Violence Survey, to include carrying of weapons both inside and outside school (2 items) | NOT INCLUDED IN QUESTIONNAIRE |
| Chronic Conditions / Illnesses | Items from the South African Demographic and Health Survey (2003) [28] the World Health Organisation Stop Tuberculosis Team, and the Kwa-Zulu Natal Department of Health TB symptom guidelines, were used to provide symptoms of pulmonary TB. Access to and adherence to TB testing and medication was measured in 4 items, based on items from the REACH (Researching Equitable Access to Healthcare) 4-province study [29].<br><br>The Health Systems Trust South African Health Review 2006 [74] was used to identify common diseases of childhood, and items from the DHS were used to identify other illnesses (16 items total). | Chronic illnesses common in South Africa such as asthma, HIV, diabetes, hypertension, low blood pressure and epilepsy will be measured using yes/no items based on clinician diagnosis.<br><br>A voluntary oral HIV-self test will be offered to participants. These have shown to have 99% sensitivity and 100% specificity [75] as well as high acceptability among users, particularly in rural areas [76].<br><br>*HIV status* will be measured using the Verbal Autopsy Questionnaire which is a checklist of AIDS-defining illnesses and AIDS-non-specific illnesses. The Verbal Autopsy uses an algorithm |

| | | |
|---|---|---|
| | *Chronic illnesses* were measured by prevalence of asthma, epilepsy and diabetes, *minor ailments* included cold/flu and worms, *acute conditions* included pneumonia, diarrhoea and burns. | of AIDS-defining illnesses to determine HIV status [77] with a specificity and sensitivity above 74% [78]. |
| Trauma | Post-traumatic stress disorder was measured using the Child PTSD Checklist (28 items) [79], accompanied by cartoons from the Levonn scale [80], which have been used in the local population [81] This was also used in our previous study of AIDS-orphanhood, and showed α.94. The Child PTSD Checklist has been used more than any other PTSD scale with Black African youth in South Africa [82,83]. On US populations, the scale shows α .82-.95. Test-retest reliability at one week was r=.91 [84]. The scale corresponds to DSM-IV diagnostic criteria. | The PTSD-8 is a short self-report instrument to screen for post-traumatic stress disorder [85]. This includes four intrusive thoughts items, two avoidance items and two hypervigilance items. These are answered on a four-point likert scale ('not at all' – 'all the time). This has not been previously used in South Africa. |
| Alcohol Use | One item from Child Behaviour Checklist [86] delinquency subscale addresses drug and alcohol use. | The 3-item Alcohol Use Disorders Identification Test Consumption (AUDIT-C) was developed as a simple method of screening for excessive drinking [87]. This measures frequency of drinking, number of units typically drunk and excessive consumption. |
| Substance Use | National survey of HIV and risk behaviour amongst young South Africans [43] (4 items) measuring multiple types of substance use. | 5 items from the Alcohol, Smoking and Substance Involvement Screening Test (ASSIST) will be used to assess substance use [88]. These cover use of tobacco products, cannabis, inhalants, nyaope |

| | | |
|---|---|---|
| | | and other drugs not prescribed by a doctor. Its validity has been studied in the South African context, yielding good results [89]. |
| Self-harm | NOT MEASURED IN EARLIER WAVES | 5 items are included to access whether the participants have ever self-injured, if yes, how old they were when they first self-injured, how many times in the last year and how long it had been since the last time and the form of self-injury. Questions are a combination of items from the Inventory of Statements About Self-Injury (ISAS) [90] and descriptive items that Prof Penelope Hasking and Assoc Professor Mark Boyes generated. The ISAS has not previously been used in South Africa. |
| Health care utilization and access | 8 Items from the REACH study – a 4-province study of health care utilization amongst HIV+ people, were used to measure health care utilization [29]. Additional open items allowed report of reasons for non-access to healthcare (this should allow estimation of accessibility, affordability and acceptability – including affordability of transport. Time spent in traveling to local clinic and mode of transport are also measured (items suggested by DoH). | 10 Items from the REACH study – a 4-province study of health care utilization amongst HIV+ people, will be used to measure health care utilization [29], traditional healers, the church or religious leaders. An additional open item will allow report of reasons for non-access to healthcare (this should allow estimation of accessibility, affordability and acceptability – including affordability of transport.<br><br>4 additional items on access and uptake of contraception have been added for women. |
| Help when you need it | NOT MEASURED IN EARLIER WAVES | 7 items will measure if the participant has a community health worker, and/or social worker and/or a religious leader and/or a traditional healer who is supporting them when they need help. If the participant yes to either one of them, a further question will ask how and in what way they support the participant. |
| Psychosis Symptoms | NOT MEASURED IN EARLIER WAVES | 6-items adapted from the Community Assessment of Psychic Experiences (CAPE) will be used [91]. This is a self-report measure for assessing phenomena that are similar to the positive symptoms of psychosis [92,93]. Each item is scored on a 4-point Likert scale, where 1= never, 2= sometimes, 3= often, 4= nearly always. |
| **YOUR CHILDHOOD** | | |

| | | |
|---|---|---|
| Your Childhood | Parental morbidity was measured at two separate stages. Firstly, in the 'Road of Life' (an adaptation of a social work tool 'the river of life' [94] children identified where parental sickness caused a change in living environment, a household move, or was a significant milestone in their lives. Secondly, children identified on the Household map whether there were any sick or disabled people in the home. Where there was a sick person, the children identified who they helped look after most, and completed a 'confidential sickness report sheet'. This sheet was placed in a separate, sealed envelope (a technique used for sensitive information in the Cape Area Panel Survey and recommended by our Child Advisory Team). The sickness report sheet identified chronicity of illness, extent of illness and frequency of illness, using items from the World Health Organisation International Classification of Functioning, Disability and Health [95] (4 items). Subscales from the 'Activity limitation and participation restriction' section (mobility and self-care domains) were used to assess extent of disability (7 items). Cause of sickness was identified using a verbal autopsy questionnaire [77], developed for use in areas with over 20% HIV-prevalence, and showing sensitivity of 83% and specificity of 75% in a Zimbabwean cohort. This questionnaire was designed primarily to identify symptoms of AIDS, and in order to a) reduce the potentially stigmatizing nature of this and b) identify other types of illness affecting caregivers, added items include the most common causes of adult illness in South Africa, as identified in the Demographic and Health Survey (2003) and the Health Systems Trust Annual Review (2006) [28,74] (18 items). | 18 items from the Adverse Childhood Experiences International Questionnaire (ACE-IQ) will measure parental morbidity, parental mortality, parental substance use, parental educational neglect, parental imprisonment, parental abandonment, violent events within the community, police violence and foster care experience [97] which has been previously used successfully with adolescents in Malawi [98]. |

| | | |
|---|---|---|
| | Parental mortality was measured at three separate stages. Firstly, in the 'Road of life' tool, and secondly in a picture activity to determine presence of multiple bereavement, dates of bereavement and cause of parental death. The qualitative picture activity was designed for use in a previous study of AIDS-orphanhood [96] and included discussion of bereavement with interviewers, and the drawing or writing of messages (children often drew flowers, or crosses, or hearts for deceased family members). Where children identified parental death, they completed a 'confidential report sheet' (which will be kept in a sealed envelope with the sickness sheet). This followed the Verbal Autopsy questionnaire [77], with added items for common alternative causes of mortality, and including non-illness causes such as road traffic accidents and homicide (18 items). The verbal autopsy questionnaire used showed sensitivity of 83% and specificity of 75% in areas of >20% HIV prevalence. | |
| Family Conflict | N/A | 2 items will assess whether there was a lot of arguing and shouting between adults at home and whether the adults at home hit each other before the participant turned 18. |
| Peer victimization / cyberbullying | Bullying was measured with the 9-item, standardised 'Social and Health Assessment Peer Victimization Scale' [48], used in research with vulnerable children in Cape Town [99] and in our previous study of AIDS-orphanhood. This scale was adapted from the Multidimensional Peer Victimization Scale, and showed α=.82 in a US validation study [100] and α=.85 in our AIDS-orphanhood study . Items include being called names, being hit or threatened and | Bullying victimization and perpetration was measured using the 12-item Brief Zurich Bullying scales (6-items victimization, 6 items perpetration) covering instances of verbal, physical and exclusionary bullying as well as sexual and homophobic events [101]. |

| | | |
|---|---|---|
| | having possessions broken or stolen. This measure generates a total global score of exposure to bullying. | |
| Violence exposure in childhood | Exposure to family conflict and domestic violence was measured using items from the UNICEF Measures for National-level monitoring of orphans and other vulnerable children [102] (2 items). Physical abuse (2 items) and emotional abuse (10 items) were measured using items from the UNICEF Measures for National-level Monitoring of Orphans and Other Vulnerable Children (Snider & Dawes, 2006). Exposure to sexual abuse was measured using three items from the Juvenile Victimization Questionnaire (JVQ) [103] and used in our previous studies in South Africa. Access to help and reactions to disclosure of abuse were measured using three items from the ISPCAN International Child Abuse Screening Tool [104]. Sense of safety in the home was measured using items from the National Primary Schools Violence Survey 2007 [105] (4 items). A further item assesses intra-household discrimination (i.e. differential allocation of food between fostered and biological children) and was developed in qualitative pilot work (1 item). | Physical, emotional and sexual child abuse exposure will be measured using the ISPCAN Child Abuse Screening Tool retrospective version [106], ICAST-R. Two additional items will measure exposure to domestic violence in the home. The tool includes four sub-scales on physical, emotional and sexual abuse and neglect with follow-up questions on when and how often this happened, which people did this and how much this experience hurt/harm them.<br><br>Access to services and reactions to abuse disclosure will be measured using adapted items from the ISPCAN Child Abuse Screening Tool – child self-report version ICAST-C [104] and WHO Multi-Country Study on Violence Against Women [40]. Questions will measure disclosure, who this disclosure was made to, whether it was supported or unsupported and any service receipt following disclosure. |
| **OVERCOMING CHALLENGES** | | |
| Self-esteem | NOT MEASURED IN EARLIER WAVES | The Rosenberg Self-Esteem Scale [107] is a widely used 10-item self-report measure for assessing individual's both negative (e.g., 'I certainly feel useless at times') and positive feelings (e.g.,'On the whole I am satisfied with myself') about themselves and their self-worth. It's a short and easy to administer scale. The items are answered using a 4-point Likert scale from strongly agree to strongly disagree. The scale has previously been used in South Africa [108]. |
| Resilience | Children may draw strength and positive outcomes from caring roles. Devised items (based on qualitative data) ask whether their caring tasks made them feel | The 12 adapted items from the Child and Youth Resilience Measure (CYRM-12), 5-point version were used to assess socio-ecological resilience among participants [109,110]. |

| | | |
|---|---|---|
| | proud or closer to the sick person (or conversely, frustrated, scared or sad). A further item asked children to identify their future goals and how they plan to achieve those goals. This gave them the opportunity to highlight the determination to become healthcare professionals. | |
| **COVID - 19** | | |
| Isolation | NOT MEASURED IN EARLIER WAVES | 7 self-developed items will ask about the participants' experience of lockdown, whom they were in lockdown and how many people, if they were in lockdown with an essential worker, if they were in lockdown at their current place and if they had access to a garden or an outside area. |
| Childcare | NOT MEASURED IN EARLIER WAVES | Two items will ask if COVID-19 had affected their employment and who cared for their children if they had to go to work. |
| Occupational Impact | NOT MEASURED IN EARLIER WAVES | 11 items will assess whether, due to COVID-19, any of the participants' household members lost their jobs, whether any of them went to bed hungry during lockdown, if anyone in the household died, if there was more violence in the household than usual, and how their mental health was during lockdown. |

3 Statistics South Africa. South AFrican Census Questionnaire 2011. 2011.

4 Statistics South Africa. General Household Survey South Africa 2019. 2020.

5 Statistics South Africa. South Africa Demographic and Health Survey 2016. 2019.

6 Consortium of Higher Education LGBT Resource Professionals. Consortium of Higher Education LGBT Resource Professionals 2018 Self-Study Report. 2018.

7 Boyden J, Dercon S. Young Lives: an international study of childhood poverty. www.younglives.org.uk 2008.

8 Promundo. IMAGES - International Men and Gender Equality Study. 2018.

9 Barnes H, Wright G. Defining child poverty in South Africa using the socially perceived necessities approach'. In: Minujin A, Nandy S, eds. *Global Child Poverty and Well-Being: Measurement, Concepts, Policy and Action*. Bristol: : Policy Press 2012. 135–54.

10 Pillay U, Roberts B, Rule S. *South African Social Attitudes. Changing Times, Diverse Voices*. Cape Town: : HSRC Press 2006.

11 Promundo. IMAGES - International Men and Gender Equality Study. 2018.

12 Labadarios D, Maunder E, Steyn N, *et al.* National food consumption survey in children aged 1-9 years: South Africa 1999. *Forum Nutr* 2003;**56**:106–9.

13 Deitchler M, Ballard T, Swindale A, *et al.* Validation of a Measure of Household Hunger for Cross-Cultural Use. Washington, DC: 2010.

14 Barnes H, Wright G. Defining child poverty in South Africa using the socially perceived necessities approach'. In: Minujin A, Nandy S, eds. *Global Child Poverty and Well-Being: Measurement, Concepts, Policy and Action*. Bristol: : Policy Press 2012. 135–54.

15 Steinert JI, Cluver LD, Meinck F, *et al.* Household economic strengthening through financial and psychosocial programming: Evidence from a field experiment in South Africa. *J Dev Econ* 2018;**134**:443–66. doi:10.1016/J.JDEVECO.2018.06.016

16 Noble M, Wright G, Cluver L. Conceptualising, defining and measuring child poverty in South Africa: An argument for a multidimensional approach. In: Dawes A, Bray R, Van der Merwe A, eds. *Monitoring child rights and wellbeing. A South African Approach*. Cape Town: : HSRC Press 2007.

17 Adolescent Pathway Project. Social support scale: Psychometric development summary. New York: : New York University 1992.

18 Van der Merwe A, Dawes A. Prosocial and antisocial tendencies in children exposed to community violence. *J Child Adolesc Ment Health* 2000;**12**:19–37.

19 Moser A, Stuck AE, Silliman RA, *et al.* The eight-item modified Medical Outcomes Study Social Support Survey: psychometric evaluation showed excellent performance. *J Clin Epidemiol* 2012;**65**:1107. doi:10.1016/J.JCLINEPI.2012.04.007

20 Sherbourne C, Stewart A. The Medical Outcomes Survey (MOS) social support survey. *Soc Sci Med* 1991;**32**:705–14.

21 Cluver L, Meinck F, Steinert J, *et al.* Parenting for Lifelong Health: A pragmatic cluster randomised controlled trial of a non-commcialised parenting programme for adolescents and their families in South Africa. *BMJ Glob Health* 2018.

22 Casale M, Wild L, Cluver L, *et al.* Social support as a protective factor for depression among women caring for children in HIV-endemic South Africa. *J Behav Med* 2015;**38**:17–27. doi:10.1007/s10865-014-9556-7

23 Berry JO, Jones WH. The parental stress scale: Initial psychometric evidence. *J Soc Pers Relatsh 1995 123 P 463-472* 1995;**12**:463–72.

24 Rochat TJ, Houle B, Stein A, *et al.* Psychological morbidity and parenting stress in mothers of primary school children by timing of acquisition of HIV infection: a longitudinal cohort study in rural South Africa. *J Dev Orig Health Dis* 2018;**9**:41–57. doi:10.1017/S204017441700068X

25 Verhoeven M, Deković M, Bodden D, *et al.* Development and initial validation of the comprehensive early childhood parenting questionnaire (CECPAQ) for parents of 1–4 year-olds. *Eur J Dev Psychol* 2017;**14**:233–47. doi:10.1080/17405629.2016.1182017

26 Elgar FJ, Waschbusch DA, Dadds MR, *et al.* Development and validation of a short form of the Alabama Parenting Questionnaire. *J Child Fam Stud* 2007;**16**:243–59.

27 Runyan D, Dunne MP, Zolotor AJ, *et al.* The development and piloting of the ISPCAN Child Abuse Screening Tool-Parent version (ICAST-P). *Child Abuse Negl* 2009;**33**:826–32. doi:10.1016/j.chiabu.2009.09.006

28 Department of Health, Medical Research Council. Demographic and Health Survey 2003. Pretoria: : Department of Health 2007.

29 Schneider H, McIntyre D. Researching Equity in Access to Health Care (REACH) - Final Technical Report. Johannesburg: 2012.

30 Health Systems Trust. South African Health Review 2009. Durban: : Health Systems Trust 2009.

31 WHO. *Measuring Health and Disability - Manual for WHO Disability Assessment Schedule WHODAS 2.0*. Geneva: : WHO 2010.

32 The Washington Group. WG Short Set on Functioning (WG-SS). 2020.https://www.washingtongroup-disability.com/question-sets/wg-short-set-on-functioning-wg-ss/ (accessed 15 Jun 2021).

33 Cluver L, Lachman J, Ward C, *et al.* Developing a parenting programme to prevent child abuse in South Africa: A pre-post pilot study. *Res Soc Work Pract* Published Online First: 2016. doi:0.1177/1049731516628647

34 Pulerwitz J, Barker G. Measuring Attitudes toward Gender Norms among Young Men in Brazil. *Men Masculinities* 2008;**10**:322–38. doi:10.1177/1097184X06298778

35 Gottert A, Barrington C, Pettifor A, *et al.* Measuring Men's Gender Norms and Gender Role Conflict/Stress in a High HIV-Prevalence South African Setting. *AIDS Behav* 2016;**20**:1785–95. doi:10.1007/s10461-016-1374-1

36 Jewkes R, Sikweyiya Y, Morrell R, *et al.* Gender Inequitable Masculinity and Sexual Entitlement in Rape Perpetration South Africa: Findings of a Cross-Sectional Study. *PLoS ONE* 2011;**6**:e29590. doi:10.1371/journal.pone.0029590

37 Pulerwitz J, Gortmaker SL, DeJong W. Measuring Sexual Relationship Power in HIV/STD Research. *Sex Roles* 2000;**42**:637–60. doi:10.1023/A:1007051506972

38 Fulu E, Jewkes R, Roselli T, *et al.* Prevalence of and factors associated with male perpetration of intimate partner violence: findings from the UN Multi-country Cross-sectional Study on Men and Violence in Asia and the Pacific. *Lancet Glob Health* 2013;**1**:e187-207. doi:10.1016/S2214-109X(13)70074-3

39 Fulu E, Miedema S, Roselli T, *et al.* Pathways between childhood trauma, intimate partner violence, and harsh parenting: findings from the UN Multi-country Study on Men and Violence in Asia and the Pacific. *Lancet Glob Health* 2017;**5**:512–22. doi:10.1016/S2214-109X(17)30103-1

40 Garcia-Moreno C, Jansen H, Ellsberg M, *et al.* WHO Multi-country Study on Women's Health and Domestic Violence against Women - Initial results on prevalence, health outcomes and women's responses. Geneva: : WHO 2005.

41 Tremblay RE, Pagani-Kurtz L, Mâsse LC, *et al.* A bimodal preventive intervention for disruptive kindergarten boys: its impact through mid-adolescence. *J Consult Clin Psychol* 1995;**63**:560–8. doi:10.1037//0022-006x.63.4.560

42 Jewkes, R, Morrell, R. Hegemonic Masculinity, Violence, and Gender Equality: Using Latent Class Analysis to Investigate the Origins and Correlates of Differences between Men. *Men Masculinities* 2018;**21**.https://journals.sagepub.com/doi/abs/10.1177/1097184X17696171 (accessed 26 Nov 2022).

43 Reproductive Health Research Unit. HIV and sexual behaviour among young South Africans: A national survey of 15-24 year olds. Parklands: : loveLife 2005.

44 Dunkle K, Jewkes R, Brown H, *et al.* Transactional sex among women in Soweto, South Africa: prevalence, risk factors and association with HIV infection. *Soc Sci Med* 2004;**59**:1581–92.

45 Medical Research Council. War at Home: Preliminary findings of the Gauteng Gender Violence Prevalence Study. Johannesburg: : Medical Research Council 2010.

46 Richters J, Martinez P. Violent Communities, family choices and children's chances: An algorithm for improving the odds. *Dev Psychopathol* 2004;**5**:609–27.

47 South African Police S, Management SS. Annual Report of the South African Police Service 2004/2005. Johannesburg: : SAPS 2005.

48 Ruchkin V, Vermeiren R, Schwab-Stone M. *The Social and Health Assessment (SAHA): Psychometric developmental summary*. New Haven: : Yale University 2004.

49 Diemer MA, McWhirter EH, Ozer EJ, *et al.* Advances in the Conceptualization and Measurement of Critical Consciousness. *Urban Rev* 2015;**47**:809–23. doi:10.1007/s11256-015-0336-7

50 Jewkes R, Nduna M, Levin J, *et al.* Impact of stepping stones on incidence of HIV and HSV-2 and sexual behaviour in rural South Africa: cluster randomised controlled trial. *BMJ* 2008;**337**:a506. doi:10.1136/bmj.a506

51 Mollica RF, Caspi-Yavin Y, Bollini P, *et al.* The Harvard Trauma Questionnaire. Validating a cross-cultural instrument for measuring torture, trauma, and posttraumatic stress disorder in Indochinese refugees. *J Nerv Ment Dis* 1992;**180**:111–6.

52 Kovacs M. Children's Depression Inventory CDI Manual. *N Y Multi-Health Syst* 1992;:1–800.

53 Suliman S. Assessing post-traumatic responses among South African Adolescents: a comparison of different methods. Cape Town: : University of Cape Town 2002.

54 Heath K, Kaminer D. *Types of Trauma exposure and severity of PTSD sypmtoms amongst Langa Adolescents*. Dep. Psychol. 2004;**MSC Thesis**.

55 Saylor C, Finch A, Spirito A. The Children's Depression Inventory: a systematic evaluation of psychometric properties. *J Consult Clin Psychol* 1984;**52**:955–67.

56 Adewuya AO, Ola BA, Afolabi OO. Validity of the patient health questionnaire (PHQ-9) as a screening tool for depression amongst Nigerian university students. *J Affect Disord* 2006;**96**:89–93. doi:10.1016/j.jad.2006.05.021

57 Bhana A, Rathod SD, Selohilwe O, *et al.* The validity of the Patient Health Questionnaire for screening depression in chronic care patients in primary health care in South Africa. *BMC Psychiatry* 2015;**15**:118. doi:10.1186/s12888-015-0503-0

58 Washington Group on Disability Statistics. An Introduction to the Washington Group on Disability Statistics Question Sets. 2020.

59 Reynolds C. Concurrent validity of What I Think and Feel: the Revised Children's Manifest Anxiety Scale. *J Consult Clin Psychol* 1980;**48**:774–5.

60 Gerard A, Reynolds C. Characteristics and applications of the Revised Children's Manifest Anxiety Scale. In: Maruish M, ed. *The use of psychological testing for treatment and planning and outcomes assessment*. Mahwah: : Lawrence Erlbaum 1999. 323–40.

61 Spitzer RL, Kroenke K, Williams JBW, *et al.* A Brief Measure for Assessing Generalized Anxiety Disorder. *Arch Intern Med* 2006;**166**:1092. doi:10.1001/archinte.166.10.1092

62 Henn C, Morgan B. Differential item functioning of the CESD-R and GAD-& in African and white working adults. *SA J Ind Psychol* 2019;**45**:10.

63 van Heyningen T, Honikman S, Tomlinson M, *et al.* Comparison of mental health screening tools for detecting antenatal depression and anxiety disorders in South African women. *PLOS ONE* 2018;**13**:e0193697. doi:10.1371/journal.pone.0193697

64 Sheehan D, Lecrubier Y, Harnett Sheehan K, *et al.* The Validity of the Mini International Neuropsychiatric Interview (MINI) according to the SCID-P and its reliabilty. *Eur Psychiatry* 1997;**12**:232–41.

65 Lecrubier Y, Sheehan D, Weiller E, *et al.* The MINI International Neuropsychiatric Interview (MINI). A short diagnostic structured interview: reliability and validity according to the CIDI. *Eur Psychiatry* 1997;**12**:224–31.

66 Horowitz LM, Bridge JA, Teach SJ, *et al.* Ask suicide-screening questions (ASQ): A brief instrument for the pediatric emergency department. *Arch Pediatr Adolesc Med* 2012;**166**:1170–6. doi:10.1001/archpediatrics.2012.1276

67 Achenbach T. Child Behaviour Checklists (CBCL/2-3 and CBCL/4-18), Teacher Report Form (TRF) and Youth Self-Report (YSR). In: Rush J, First M, Blacker D, eds. *The Handbook of Psychiatric Measures*. Arlington, VA: : The American Psychiatric Association 2000.

68 Barbarin OA, Richter L, deWet T. Exposure to violence, coping resources, and psychological adjustment of South African children. *Am J Orthopsychiatry* 2001;**71**:16–25. doi:10.1037/0002-9432.71.1.16

69 Achenbach T, Rescorla L. Manual for the ASEBA School-Age Forms and Profiles. Burlington, VT: : University of Vermont 2001.

70 Song L-Y, Singh J, Singer M. The Youth Self-Report Inventory: A study of its measurement fidelity. *Psychol Assess* 1994;**6**:245–326.

71 Goodman R. The Strengths and Difficulties Questionnaire: A research note. *J Child Psychol Psychiatry* 1997;**38**:581–6.

72 Goodman R, Cluver L, Tshandu V, *et al.* Xhosa version of the Strengths and Difficulties Questionnaire (Child report and Parent report). London: : Institute of Psychiatry 2004.

73 Goodman R. Psychometric Properties of the Strengths and Difficulties Questionnaire (SDQ). *J Am Acad Child Adolesc Psychiatry* 2001;**40**:1337–45.

74 Ijumba P, Padarath A, Health Systems Trust. South African Health Review 2006. Durban: : Health Systems Trust 2006.

75 Devillé W, Tempelman H. Feasibility and robustness of an oral HIV self-test in a rural community in South-Africa: An observational diagnostic study. *PLOS ONE* 2019;**14**:e0215353. doi:10.1371/journal.pone.0215353

76 Hector J, Davies M-A, Dekker-Boersema J, *et al.* Acceptability and performance of a directly assisted oral HIV self-testing intervention in adolescents in rural Mozambique. *PloS One* 2018;**13**:e0195391. doi:10.1371/journal.pone.0195391

77 Lopman B, Barnabas R, Boerma T, *et al.* Creating and validating an algorithm to measure AIDS mortality in the adult population using verbal autopsy. *Public Libr Sci Med* 2006;**3**:e312.

78 Lopman B, Cook A, Smith J, *et al.* Verbal autopsy can consistently measure AIDS mortality: a validation study in Tanzania and Zimbabwe. *J Epidemiol Community Health* 2010;**64**:330–4. doi:10.1136/jech.2008.081554

79 Amaya-Jackson L. Child PTSD Checklist. North Carolina: : Duke Treatment Service 1995.

80 Richters J, Martinez P, Valla J. Levonn: A cartoon-based interview for assessing children's distress symptoms. Child and Adolescent Disorders Research Branch Division of Clinical Research National Institute of Mental Health 5600 Fishers Lane, Room 18c-17 Rockville, maryland 20857 tel (301) 443-5944: : University of Maryland, NIMH 1990.

81 Zissis C, Ensink K, Robertson B. A Community Study of taxi violence and distress symptoms among youth. *J Child Adolesc Ment Health South Afr* 2000;**12**:151–61.

82 Seedat S, Nyamai C, Njenga F, *et al.* Trauma Exposure and Post-Traumatic Stress symptoms in urban African schools. *Br J Psychiatry* 2004;**184**:169–75.

83 Amaya-Jackson L, Newman E, Lipschitz D. The Child and Adolescent PTSD Checklist in Three Clinical Research Populations. New York: : Annual Meeting of the American Academy of Child and Adolescent Psychiatry 2000.

84 Newman E, Amaya-Jackson L. Assessment of trauma instruments for children. In: *The Scientific Proceedings of the 12th International Conference for Traumatic Stress Studies*. San Fransisco: 1996.

85 Hansen M, Andersen TE, Armour C, *et al.* PTSD-8: A Short PTSD Inventory. *Clin Pract Epidemiol Ment Health* 2010;**1**:101–8. doi:10.2174/1745017901006010101

86 Achenbach T. Manual for the Child Behaviour Checklist/2-3 and 1992 Profile. Burlington, VT: : University of Vermont 1992.

87 Saunders JB, Aasland OG, Babor TF, *et al.* Development of the Alcohol-Use Disorders Identification Test (Audit) - Who Collaborative Project on Early Detection of Persons with Harmful Alcohol-Consumption .2. *Addiction* 1993;**88**:791–804. doi:10.1111/j.1360-0443.1993.tb02093.x

88 WHO ASSIST Working Group. The Alcohol, Smoking and Substance Involvement Screening Test (ASSIST): development, reliability and feasibility. *Addiction* 2002;**97**:1183–94.

89 van der Westhuizen C, Wyatt G, Williams JK, *et al.* Validation of the Alcohol, Smoking and Substance Involvement Screening Test in a low- and middle-income country cross-sectional emergency centre study. *Drug Alcohol Rev* 2016;**35**:702–9. doi:10.1111/dar.12424

90 Klonsky ED, Glenn CG. Assessing the functions of non-suicidal self-injury: Psychometric properties of the Inventory of Statements about Self-Injury (ISAS). *J Psychopathol Behav Assessment2* 2009;**31**:215–9.

91 Aloba O, Opakunle T. The Brief 10-Item Community Assessment of Psychic Experiences-Positive Scale (Brief CAPE-P10): Initial psychometric properties, gender measurement invariance and mean differences among Nigerian adolescents. *Early Interv Psychiatry* 2020;**14**:723–33. doi:10.1111/EIP.12903

92 Ribeaud D, Murray A, Shanahan L, *et al.* Cohort Profile: The Zurich Project on the Social Development from Childhood to Adulthood (z-proso). *J Dev Life-Course Criminol* 2022;**8**:151–71. doi:10.1007/s40865-022-00195-x

93 Shanahan L, Steinhoff A, Bechtiger L, *et al.* Frequent teenage cannabis use: Prevalence across adolescence and associations with young adult psychopathology and functional well-being in an urban cohort. *Drug Alcohol Depend* 2021;**228**:109063. doi:10.1016/j.drugalcdep.2021.109063

94 Buchanan A. *Social work tools for assessment of children*. Oxford: : Oxford University 2002.

95 World Health Organization. ICF Checklist Version 2.1a, Clinician Form for International Classification of Functioning, Disability and Health. Geneva: : WHO 2003.

96 Cluver L, Gardner F, Operario D. Psychological distress amongst AIDS-orphaned children in urban South Africa. *J Child Psychol Psychiatry* 2007;**48**:755–63. doi:10.1111/j.1469-7610.2007.01757.x

97 World Health Organization. Adverse Childhood Experiences International Questionnaire (ACE-IQ) - Rationale for ACE-IQ. Geneva: : WHO 2012. http://www.who.int/violence_injury_prevention/violence/activities/adverse_childhood_experiences/en/

98 Kidman R, Smith D, Piccolo LR, *et al.* Psychometric evaluation of the Adverse Childhood Experience International Questionnaire (ACE-IQ) in Malawian adolescents. *Child Abuse Negl* 2019;**92**:139–45. doi:10.1016/J.CHIABU.2019.03.015

99 Ward CL, Martin E, Theron C, *et al.* Factors affecting resilience in children exposed to violence. *South Afr J Psychol* 2007;**37**:164–87.

100 Mynard H, Joseph S. Development of the Multidimensional Peer-Victimization Scale. *Aggress Behav* 2000;**26**.

101 Murray AL, Eisner M, Ribeaud D, *et al.* Validation of a Brief Self-Report Measure of Adolescent Bullying Perpetration and Victimization. *Assessment* 2021;**28**:128–40. doi:10.1177/1073191119858406

102 Snider L, Dawes A. Psychosocial Vulnerability and Resilience Measures For National-Level Monitoring of Orphans and Other Vulnerable Children: Recommendations for Revision of the UNICEF Psychological Indicator. Cape Town: : UNICEF 2006.

103 Finkelhor D, Hamby SL, Ormrod R, *et al.* The Juvenile Victimization Questionnaire: Reliability, validity, and national norms. *Child Abuse Negl* 2005;**29**:383–412. doi:10.1016/j.chiabu.2004.11.001

104 Zolotor AJ, Runyan DK, Dunne MP, *et al.* ISPCAN Child Abuse Screening Tool Children's Version (ICAST-C): Instrument development and multi-national pilot testing. *Child Abuse Negl* 2009;**33**:833–41. doi:http://dx.doi.org/10.1016/j.chiabu.2009.09.004

105 Burton P. National Primary School Violence Survey 2007. Cape Town: : Centre for Justice and Crime Prevention 2008.

106 Dunne MP, Zolotor AJ, Runyan DK, *et al.* ISPCAN Child Abuse Screening Tools Retrospective version (ICAST-R): Delphi study and field testing in seven countries. *Child Abuse Negl* 2009;**33**:826–32. doi:10.1016/j.chiabu.2009.09.005

107 Rosenberg M. Society and the adolescent self-image. *Princet NJ Princet Univeristy Press* 1965.

108 Sherr L, Macedo A, Tomlinson M, *et al.* A foot in the door: A report on the Child Community Care study evaluating the effect of community-based organisation support on child wellbeing in HIV-affected communities. University College London 2016.

109 Liebenberg L, Ungar M, LeBlanc JC. The CYRM-12: a brief measure of resilience. *Can J Public Health Rev Can Sante Publique* 2013;**104**. doi:10.1007/BF03405676

110 Ungar M, Liebenberg L. Assessing Resilience Across Cultures Using Mixed Methods: Construction of the Child and Youth Resilience Measure. *J Mix Methods Res* 2011;**5**:126–49. doi:10.1177/1558689811400607
