## Supplement 2 for "Interrupting the intergenerational cycle of violence: protocol for a three-generational longitudinal mixed-methods study in South Africa (INTERRUPT_VIOLENCE)"

Supplement 2: Young Carers 3 – Caregiver Questionnaire

| ABOUT YOU | |
|---|---|
| Demographic information | Age, home language and basic demographic will be asked using four items [1], South Africa General Household Survey [2], and the South Africa Demographic and Health Survey [3]. Questions on sexuality, gender and sex assigned at birth were modeled on the US Consortium of Higher Education LGBT Resource Professionals Self-Study [4]. |
| Education | Highest level of education attained will be measured using items from the IMAGES Study South Africa 2018 [5].<br><br>Additional items will focus on completed schooling: school dropout and reasons for drop out and whether they accessed tertiary education.<br><br>Violence exposure at school was measured with one item. |
| Your status | 5 items will ask about their citizenship status, whether they are a South African citizen, if no, which country, if they have a birth certificate, ID book or passport and if no, why they don't have. |
| **YOUR HOME** | |
| Your home | 5 items will identify whether the caregiver lives with the young adult and the type of accommodation they currently live in vs the type of accommodation they occupied 10 years ago at study follow-up.<br><br>21 items will be used to identify size of the home, type of home, number of people living in it and the relationships to the participants based on items from the South African Census [1] and IMAGES Study [5]. These cover if house is owned or rented, number of rooms, kitchen, toilets, number of chronically ill people in the home, number of employed people, whether adults and children share beds, the household water and electricity source and how many days either were not available in the past month. |
| Employment | Six items from the IMAGES Study South Africa [6] assess employment and household income focusing on employment of respondent, whether the work is seasonal, part-time or full-time, the monthly household income, number of adults and children dependent on the household income. |
| Making ends meet | Food insecurity will be measured using the three item Household Hunger Scale (HHS). This is a subscale of the HFIAS (Household Food Insecurity Scale). It consists of 3 items and has demonstrated the potential for cross-cultural validity. The HFIAS has been validated throughout different countries, including South Africa [7]. One additional item is included to assess whether children had enough food in the past 4 weeks.<br><br>8 items from necessities approach of Barnes and Wright [8] and 6 additional items will measure what necessities the household was able to afford in the past month. These will include three meals a day, sanitary products, household cleaning products, electricity, water, shoes amongst others. |

| | |
|---|---|
| | 4-items from the Sinovuyo Teen Study will assess a household's financial management including membership in e.g. a stokvel or burial society, borrowing money and ability to cope with financial shocks [9]. |
| Government grants | Receipt within the household of the major forms of social security transfers (Child Support Grant, Foster Child Grant, Care Dependency Grant, Disability Grant, Grant in Aid, government housing subsidy, Old Age Pension) will be measured. An additional item will ask if the household received the special COVID-19 Social Relief of Distress grant and the one time R700 grant provided in 2020 as a result of the COVID-19 pandemic. |
| Communicating with others | Two items will assess whether households have access to a mobile phone or smart phone, TV, radio and computer/internet and whether and what type of health messaging they may have received through their devices in the past year. |
| **YOUR FAMILY AND FRIENDS** | |
| Social Support | The 8-item Modified Medical Outcome Study Social Support Survey (mMOS-SS) will be used to measure social support [10] which has similarly excellent validity compared to the original 18-item MOS-SS. The MOS-SS was designed to measure social support received by persons with chronic conditions. It has good internal consistency α=0.91to 0.97 and test-retest reliability α=0.72 to 0.78 [11]. It has been used in our previous studies in South Africa with good success [12,13]. |
| **YOUR CHILDREN** | |
| Your relationship with (name of adult participant) | 9 items will be asked to assess the relationship between the participants and the young adult participating in the study, whether the young adult lived with the participant for the whole duration of her/his childhood, if the young adult is biological, whether she/he breastfed, if the participants still financially supporting the young adult and how it was to care for the young adult. |
| Disciplining and Protecting (name of young adult participant) | The ISPCAN Child Abuse Screening Tool – ICAST-P (parent self-report) will be used to on the use of physical and emotional punishment and the young adults' experience of neglect, sexual abuse, domestic violence and sibling bullying. Items on positive discipline are also included [14]. This will be adapted to allow retrospective reporting of abuse when caregivers were parenting the young adult during their adolescence. |
| Illness in early childhood (young adult) | One item will assess whether the caregiver had to take the young adult to a hospital during their childhood for illnesses such as pneumonia, HIV, malnutrition, burns, convulsions, diarrhoea or injuries. |
| Current children you are caring for | 14 items will assess whether the caregiver currently has a child in their care, their relationship to that child and the roles of caregiving they carry out. These will also assess whether the caregiver had children that have died and the causes they have died of. |

| | |
|---|---|
| Pregnancy | Two items for women assess the number of times they have been pregnant and whether the pregnancy for the last child they are caring for was planned. |
| Child protection | Two items will assess whether the child are currently caring for was exclusively breastfed for 6 months, and whether their birth was registered |
| Fathering section | For men, one item will assess whether the pregnancy had been planned. |
| Parenting stress | Parenting stress will be assessed using the 18-item Parenting Stress Scale (PSS) which was developed as an alternative to the 101-item Parenting Stress Index [15]. It provides a measure that considers positive aspects of parenting as well as the negative, focusing on traditional "stressful" aspects. The PSS has previously been used in a longitudinal cohort study in South Africa [16]. |
| Children's education | Nine items will assess whether the index child attends crèche or school, whether they receive a fee exemption, receive free school books, school meals, school trips, school uniform or school transport. |
| Early childhood parenting | 38 items from the Comprehensive Early Childhood Parenting Questionnaire (CECPAQ) support, structure and stimulation sub-scales will be used to assess engagement with the index child, creating play opportunities for the index child, disciplining the child and consistency of parenting behaviours [17]. This measure has not been previously used in South Africa but was piloted extensively with some cultural adaptations e.g. toys and games specific to South Africa.<br><br>7 items from Involved Parenting subscale of the Alabama Parenting Questionnaire [18] measured parental involvement in the index child's activity. These have been previously used in South Africa with good validity and reliability [12]. |
| Attitudes to corporal punishment | Three self-developed items assessed attitudes towards disciplining children. These cover endorsement of physical discipline, gendered discipline and parenting similarly vs differently to one's own parents. |
| Child abuse and neglect – Disciplining child | 39 items from the ISPCAN Child Abuse Screening Tool parent self-report (ICAST-P) will be used to self-report on the use of physical and emotional punishment and the child' experience of neglect, sexual abuse, domestic violence and sibling bullying. Items on positive discipline are also included [14]. |
| The health of your children - oldest child in your care | Seven items covering injuries and burns, diarrhea and vomiting, pneumonia or bronchitis, skin conditions, general body pains, cold/flu and COVID-19 will be retained from the previous questionnaire. Pulmonary TB will be measured using the TB symptom assessment used in the baseline and follow-up research. Access to health care will be assessed with 8 items from the REACH study [19].<br><br>Disability will be assessed using 9 items from the Short Set on Functioning Washington Group Questionnaire (WG-SS) [20]. These cover visual, auditory, mobility and cognitive disabilities. |
| HIV - oldest child in your care | 9 items adapted from Sinovuyo Teen Study (2015) will be asked to assess if their children have been tested for HIV, if yes, are they positive, if yes, are they taking any medication, did they need to go to hospital in the last month, |

| | |
|---|---|
| | did they tell their child that they are HIV-positive, do they know their child's CD4 and if they can tell us [21] |
| **YOUR RELATIONSHIPS** | |
| Your relationships when (adult participant) was growing up | Participants' relationship status and experiences will be measured using items from the IMAGES Study South Africa 2018 [5]. Items will assess current partners and partners 10 years ago when the adolescent lived with them. The items will assess what their main motivation in staying in the relationship if they have one, if their partners drink alcohol, use drugs or have HIV, how many one night stands they had in the past year and if any of their sexual partners were from the same sex. These items also assess the nature of the relationships ranging from main partners to casual partners and side partners. |
| Relationship Beliefs (male & female version) | The Gender Equitable Men (GEM) Scale measures attitudes toward gender norms in intimate relationships or differing social expectations for men and women [22]. 8 items from the Inequitable scale will be used and 2 additional items specific to the South African context on lobola and rape were added. Each item is scored on a 3-point scale, where 1 = agree, 2 = partially agree, and 3 = do not agree. The estimate of internal consistency = .81, but it is expected to vary according to the setting. This measures has been previously used in South Africa [23,24]. |
| Relationship control scale (male & female version) | The Sexual Relationship Power Scale (SRPS) measures power within sexual relationships for both men and women. 8 items of the Relationship Control subscale are used in the questionnaire. Items are scored on a 4-point Likert scale, with 1 = strongly agree, 2 = agree, 3 = disagree, and 4 = strongly disagree) [25]. |
| Intimate Partner Violence | IPV perpetration will be measured using the 19-item questionnaire from the UN Multi-country cross-sectional study on men and violence and Asia and the Pacific [26,27]. Questions will ask if the behaviour happened in the past 12 months, if yes, the frequency in the last year and the frequency in the woman's lifetime.<br><br>IPV experience will be measured using the 25 item WHO domestic violence instrument from the WHO Multi-Country Study on Violence Against Women [28] for past year and lifetime exposure. Five additional items assess IPV experience in pregnancy. For women, additional questions cover whether they have sought help, whom they have sought help from and what the response to their disclosure was. |
| Community Violence | 8 items from WHO community violence perpetration instrument developed by Tremblay et al., [29] and adapted by Jewkes & Morrell [30] will be used to assess whether a participant has taken part in violent activities within the community in which they live . |
| Risky sex | Items from the National survey of HIV and risk behaviour amongst young South Africans [31] will be retained to measure sexual activity (screening item), age of sexual debut, frequency of condom use, sex with an older partner, sexual activity under the influence of alcohol and under the influence of drugs, and |

| | |
|---|---|
| | of difficulty, and 3 = cannot do at all. The WG-SS has been extensively tested and validated and used in over 80 countries [40]. |
| Anxiety | The General Anxiety Disorder (GAD)-7 seven item questionnaire measures symptoms of GAD following DSMV-IV symptom criteria for GAD and assigns scores of 0, 1, 2, and 3, to the response categories of 'not at all', 'several days', 'more than half the days', and 'nearly every day', respectively. Using the threshold score of 10, the GAD-7 has a sensitivity of 89% and a specificity of 82% for GAD [41]. This tool and its 2-item form have been used in South Africa [42,43] |
| Suicide Screening Questions | 5 items are from the National Institute for Mental Health's Ask Suicide Screening Questions (ASQ) Toolkit [44]. It was developed as a brief, straight-forward tool to identify potential suicide risk. It includes items like 'in the past few weeks, have you wished you were dead' and 'have you ever tried to kill yourself'. |
| Chronic conditions / illnesses | HIV status will be measured using the Verbal Autopsy Questionnaire which is a checklist of AIDS-defining illnesses and AIDS-non-specific illnesses. The Verbal Autopsy uses an algorithm of AIDS-defining illnesses to determine HIV status [45] with a specificity and sensitivity above 74% [46].<br><br>A voluntary oral HIV-self test will also be offered to participants. These have shown to have 99% sensitivity and 100% specificity [47] as well as high acceptability among users [48].<br><br>Chronic illnesses common in South Africa such as asthma, HIV, diabetes, hypertension, low blood pressure and epilepsy will be measured based on clinical diagnosis. |
| Trauma | The PTSD-8 is a short self-report instrument to screen for post-traumatic stress disorder [49]. This includes four intrusive thoughts items, two avoidance items and two hypervigilance items. These are answered on a four-point likert scale ('not at all' – 'all the time). This has not been previously used in South Africa. |
| Alcohol Use | The 3-item Alcohol Use Disorders Identification Test Consumption (AUDIT-C) was developed as a simple method of screening for excessive drinking **[50]**. This measures frequency of drinking, number of units typically drunk and excessive consumption. |
| Substance Use | 5 items from the Alcohol, Smoking and Substance Involvement Screening Test (ASSIST) will be used to assess substance use **[51]**. These cover use of tobacco products, cannabis, inhalants, nyaope and other drugs not prescribed by a doctor. Its validity has been studied in the South African context, yielding good results **[52]**. |
| Accessing health services | 10 Items from the REACH study – a 4-province study of health care utilization amongst HIV+ people, will be used to measure health care utilization [19], traditional healers, the church or religious leaders. An additional open item will allow report of reasons for non-access to healthcare (this should allow estimation of accessibility, affordability and acceptability – including affordability of transport. |

| | |
|---|---|
| | 4 additional items on access and uptake of contraception have been added for women. |
| Help when you need it | 7 items will measure if the participant has a community health worker, and/or social worker and/or a religious leader and/or a traditional healer who is supporting them when they need help. If the participant yes to either one of them, a further question will ask how and in what way they support the participant. |
| Psychotic experiences | 6-items adapted from the Community Assessment of Psychic Experiences (CAPE) will be used [53]. This is a self-report measure for assessing phenomena that are similar to the positive symptoms of psychosis [54,55]. Each item is scored on a 4-point Likert scale, where 1= never, 2= sometimes, 3= often, 4= nearly always. |
| **YOUR CHILDHOOD** | |
| Your childhood<br><br>Child abuse and neglect | Physical, emotional and sexual child abuse exposure will be measured using the ISPCAN Child Abuse Screening Tool retrospective version [56], ICAST-R. Two additional items will measure exposure to domestic violence in the home. The tool includes four sub-scales on physical, emotional and sexual abuse and neglect with follow-up questions on when and how often this happened, which people did this and how much this experience hurt/harm them.<br><br>Access to services and reactions to abuse disclosure will be measured using adapted items from the ISPCAN Child Abuse Screening Tool – child self-report version ICAST-C [57] and WHO Multi-Country Study on Violence Against Women [28]. Questions will measure disclosure, who this disclosure was made to, whether it was supported or unsupported and any service receipt following disclosure. |
| Family conflict | 2 items will assess whether there was a lot of arguing and shouting between adults at home and whether the adults at home hit each other before the participant turned 18. |
| Adverse life events | 18 items from the Adverse Childhood Experiences International Questionnaire (ACE-IQ) will measure parental morbidity, parental mortality, parental substance use, parental educational neglect, parental imprisonment, parental abandonment, violent events within the community, police violence and foster care experience [58] which has been previously used successfully with adolescents in Malawi [59]. |
| Peer victimization | Bullying victimization and perpetration was measured using the 12-item Brief Zurich Bullying scales (6-items victimization, 6 items perpetration) covering instances of verbal, physical and exclusionary bullying as well as sexual and homophobic events [60]. |
| **OVERCOMING CHALLENGES** | |
| Self-esteem | The Rosenberg Self-Esteem Scale [61] is a widely used 10-item self-report measure for assessing individual's both negative (e.g., 'I certainly feel useless |

| | |
|---|---|
| | at times') and positive feelings (e.g.,'On the whole I am satisfied with myself') about themselves and their self-worth. It's a short and easy to administer scale. The items are answered using a 4-point Likert scale from strongly agree to strongly disagree. |
| Resilience | The 12 adapted items from the Child and Youth Resilience Measure (CYRM-12), 5-point version were used to assess socio-ecological resilience among participants [62,63]. |
| **COVID-19** | |
| COVID-10 Isolation | 7 self-developed items will ask about the participants' experience of lockdown, whom they were in lockdown and how many people, if they were in lockdown with an essential worker, if they were in lockdown at their current place and if they had access to a garden or an outside area. |
| Childcare | Two items will ask if COVID-19 had affected their employment and who cared for their children if they had to go to work. |
| Occupational Impact (Employment) | 11 items will assess whether, due to COVID-19, any of the participants' household members lost their jobs, whether any of them went to bed hungry during lockdown, if anyone in the household died, if there was more violence in the household than usual, and how their mental health was during lockdown. |

**References**


1 Statistics South Africa. South AFrican Census Questionnaire 2011. 2011.

2 Statistics South Africa. General Household Survey South Africa 2019. 2020.

3 Statistics South Africa. South Africa Demographic and Health Survey 2016. 2019.

4 Consortium of Higher Education LGBT Resource Professionals. Consortium of Higher Education LGBT Resource Professionals 2018 Self-Study Report. 2018.

5 Promundo. IMAGES - International Men and Gender Equality Study. 2018.
