## Supplement 3 for "Interrupting the intergenerational cycle of violence: protocol for a three-generational longitudinal mixed-methods study in South Africa (INTERRUPT_VIOLENCE)"

Supplement 3: Young Carers 3 – Young Children Questionnaire

| ABOUT ME | |
|---|---|
| Demographic information | Name, age, gender will be asked about using items from the Child Community Care Study [1] . |
| Early Childhood Education | This will be measured with two items asking if the child attends nursery/ or if of school age attended nursery.<br><br>If child is of school age, three items will measure if children attend school, if they had to stay off school to attend to household chores and how often this happened.<br><br>Further they are asked if they have fun at school/crèche, whether they have friends and if they feel safe at school/crèche.<br><br>For children in school, 2 items measure after school activities offered by the school and current grade. |
| Feeling faces game | The aim of this 'game' is to help participants understand the basic feelings of happy, sad, scared and angry in the form of faces. Understanding these feelings is comprised of being able to name them, recognise them for oneself and recognize them in others. It provides a common understanding regarding some of the measures in the questionnaire that incorporate feeling words. The game is also intended to be fun and playful and help with engaging child participants in the research. |
| **MY HOME AND ME** | |
| Home environment | This will be measured using seven items from the Mad About ART study [2]. One item will measure number of people living in the home. Six items will measure if the child helps to look after younger children at home, helps to look after unwell people at home, sleeps in their own bed, lives with someone who has a job, has a dry and warm home, sleeps in their own bed. |
| People in the home | This will be assessed using the House and Community Plan [3,4] used in forensic social work. The child draws their house and community and then makes play-doh figures for the people staying in the house. The technique's goal is to focus on the child's daily movements from one place to another. With this visual and interactive play-related communication technique, it is possible to identify places where a child feels secure and identify areas and situations where they feel threatened. Through this technique, a child is also allowed to express both positive and negative emotions. Children are asked to talk about the different rooms in their home, the different people staying in the rooms and then also the outside community places that children engage with e.g. school, youth centre, shops etc. |
| **MY CAREGIVER AND ME** | |

| | |
|---|---|
| Caregiver Status | The relationship of the child to primary caregiver will be measured. Primary caregiver will be identified as the person who 'stays with you and takes care of you at home'. Relationships of caregiver to children will be categorized as the following: mother, father, brother, sister, aunt, uncle, grandmother, grandfather and other. If children do not live with their parent, three follow-up items will query where their parents are now, whether they have died and who else in the family home has died during the past two years. |
| Parenting and Home Environment | 6 items from the Child Community Care Study will be used measuring feelings of belonging, treatment equal to other children in the home, and whether the child has recently changed caregiver.<br><br>Parenting will be measured using 11 adapted items from the Comprehensive Early Childhood Parenting Questionnaire (CECPAQ) focusing on parental involvement, positive parenting, harsh and abusive parenting [5]. |
| Negative Family Processes | 4-items adapted from the ICAST-C [6] assess children's witnessing of domestic violence between adults. Three additional items assess whether children are scared by the fighting, whether they think it is their fault and whether they have disclosed this to anyone.<br><br>Children's attitudes and beliefs about the acceptability of family violence are measured using 7 items from the Attitudes About Family Violence (AAFV) scale [7]. The child is asked to rate these using a using a 3-point (adapted from 5-point Likert-type scale) on how much each statement reflects the child's beliefs. Scale anchors are "Always, sometimes, never". |
| **OTHER CHILDREN AND ME** | |
| Friendships | Two items will measure whether the child has a friend and the activities they do together. |
| Peer Bullying & Sibling Bullying | Bullying will be measured with the 9-item, standardized 'Social and Health Assessment Peer Victimization Scale' [8], used in research with vulnerable children in Cape Town [9]. This scale was adapted from the Multidimensional Peer Victimization Scale, which was validated in the US [10]. Items include: being called names, being hit or threatened and having possessions broken or stolen, being hurt, being stood too close to. This measure generates a total global score of exposure to bullying.<br>Five additional items ask about bystander intervention, possible disclosure of bullying and outcomes of that disclosure. |
| **MY HEALTH AND FOOD** | |
| Food Security | Food insecurity will be measured using an item from the South African National Food Consumption Survey (1999) [11] asking if the child went to bed hungry the previous night. |

| | |
|---|---|
| Physical Health | Physical health will be measured through a combination of tools. Common childhood illnesses such as diarrhoea, bilharzia or colds will be measured using 8 items. 5 items will measure physical disabilities such as problems seeing, hearing or walking/running adapted from the Washington Group Short Set on Functioning [12].<br><br>Child height and weight will be taken using a measuring tape and scales. |
| Medical care | 11 items adapted from the REACH Study will about "Seeing people for your health": frequency of clinic visits, reasons for clinic visits, pills taken regularly, hospital stays and reasons, anyone in family sick, any in the family taking pills regularly [13]. |
| Family HIV and Child HIV status | These will be measured using three items from the MAD about ART study [2]. |
| Post-Traumatic Stress Disorder | PTSD will be measured using the 10 item Trauma Symptoms Checklist for Young Children [14]. This measures the frequency of symptoms of post-traumatic stress and has previously been used in the Child Community Care Study with children of similar ages in South Africa, Malawi and Zambia [1]. One item was added to measure how scared participants feel. |
| Child Anxiety Symptoms | Anxiety will be measured using the Revised Children's Manifest Anxiety Scale with 14 items. In the previous orphan study, the full scale showed an α.80 (2005), and the reduced scale showed α .75 [15] and .80 [16]. The RCMAS has been standardized in US populations and has previously been used in poor urban communities in Cape Town [17]. |
| Suicide Ideation Symptoms | Mini International Psychiatric Interview for Children and Adolescents suicidality and self-harm subscale (5 items) [18]. The MINI-Kid has been extensively validated in developed world populations and shows strong internal consistency and test-retest reliability. The MINI-kid has previously been used in our previous studies in South Africa [19]. |
| Child Depressive Symptoms | The Child Depression Inventory- Short Form (10 items) [20], was used in our previous studies of AIDS-orphanhood and showed an acceptable α=.67 [21] and α=.69 [22]. The CDI has been used in multiple South African populations [23], including an adapted version, which was validated against the Beck Depression Inventory (r=0.81).<br><br>A four additional items assess if they get in trouble a lot, if they feel sad a lot, what makes them feel better, and what adults can do when children feel sad, scared, or worried. |
| **MY COMMUNITY** | |
| Safe Spaces and Support | 16-items self-developed items assess whether the child feels safe or unsafe in their community, someone they can talk to when they need help, and how they feel in their community |

| | |
|---|---|
| Community Violence | 4-items from the Things I Have Seen and Heard Scale assess how often they have been attacked outside their home, seen someone stabbed/beaten/shot, been sexually assaulted, or had something stolen [24] |
| Behavior Problems | The Strengths and Difficulties Questionnaire (SDQ) - Conduct Problems Subscale (5 items) is used to assess behavior problems [25]. The SDQ is well-validated, and has been translated into 51 languages, including isiXhosa and isiZulu. |
| Resilience | The Brief Child and Youth Resilience Measure (CYRM-12) is a measure of the resources (individual, relational, communal and cultural) available to individuals that may bolster their resilience [26]. The measure has 3 subscales accounting for personal, relational, and contextual factors implicated in resilience processes. It was originally designed for use with youth aged 9 to 23 years old. The measure has 12 items and a 3-point response scale. The CYRM has been validated in South Africa [27]. |
| Distress | One item will assess how upset/distressed the child was by the questions asked in the questionnaire<br><br>A section for interviewers will require detailed case notes and reflections about each child interview. |
